# Towards 3D Deep Learning for neuropsychiatry: predicting Autism diagnosis using an interpretable Deep Learning pipeline applied to minimally processed structural MRI data

**DOI:** 10.1101/2022.10.18.22281196

**Authors:** Mélanie Garcia, Clare Kelly

## Abstract

By capitalizing on the power of multivariate analyses of large datasets, predictive modeling approaches are enabling progress toward robust and reproducible brain-based markers of neuropsychiatric conditions. While Deep Learning offers a particularly promising avenue to further advance progress, there are challenges related to implementation in 3D (best for MRI) and interpretability. Here, we address these challenges and describe an interpretable predictive pipeline for inferring Autism diagnosis using 3D Deep Learning applied to minimally processed structural MRI scans. We trained 3D Deep Learning models to predict Autism diagnosis using the openly available ABIDE I and II datasets (n = 1329, split into training, validation, and test sets). Importantly, we did not perform transformation to template space, to reduce bias and maximize sensitivity to structural alterations associated with Autism. Our models attained predictive accuracies equivalent to those of previous Machine Learning studies, while side-stepping the time- and resource-demanding requirement to first normalize data to a template, thus minimizing the time required to generate predictions. Further, our interpretation step, which identified brain regions that contributed most to accurate inference, revealed regional Autism-related alterations that were highly consistent with the literature, such as in a left-lateralized network of regions supporting language processing. We have openly shared our code and models to enable further progress towards remaining challenges, such as the clinical heterogeneity of Autism, and to enable the extension of our method to other neuropsychiatric conditions.

## 2 Introduction

Autism Spectrum Disorder (Autism) is a complex and heterogeneous neurodevelopmental condition characterized by divergence from typical development on a number of behavioral dimensions, including communication, social interaction, and repetitive or restricted behaviors or areas of interest[1]. These manifest behaviors likely reflect developmental neurological alterations over the lifespan[2-7], a suggestion supported by structural MRI studies[2, 8-26]. Despite substantial research effort, however, no compelling brain-based biomarkers have yet emerged. Autism Spectrum Disorder is diagnosed through clinician judgment and gold standard observational tests, such as the Autism Diagnostic Observation Schedule (ADOS)[27] and the Autism Diagnostic Interview-Revised (ADI-R)[28], typically around age 43 months[29]. Given the considerable heterogeneity inherent to the diagnosis, and the wide range of long-term outcomes, the availability of robust and reproducible brain biomarkers for Autism could help refine diagnoses and treatment plans, thus promoting better outcomes. The availability of predictive models could also help clinicians build personalized care paths[30].

One challenge in the search for biomarkers and in the development of predictive models is the attainment of sample sizes that afford adequate statistical power. This challenge is exacerbated by clinical heterogeneity[30]. Multi-site collaborative studies yielding well-powered samples, such as ABIDE I and II[31-32], have gone some way to addressing this challenge, and analyses of these samples suggest a distributed pattern of Autism-related structural alterations[16-18, 20, 21, 24, 33-34]. The application of multivariate approaches, such as Machine and Deep Learning, offer another promising avenue for the search for brain-based biomarkers and the construction of predictive models.

These methods enable the simultaneous exploration of a very large set of features, offering much more powerful analytical capacity than univariate approaches. To date, such approaches have had moderate success, with recently reported prediction accuracies (for Autism diagnosis) in the range of 65-70% for models built using both functional and structural MRI data[35-38]. In an effort to boost accuracy through competition, Traut et al.[39] held an international challenge in which competing teams predicted Autism diagnosis using a large multisite dataset comprising preprocessed anatomical and functional MRI data from > 2,000 individuals. Of the 589 models submitted, the 10 best were combined and evaluated using a subset of unseen data (from one of the sites included in the main dataset), as well as data from an additional, independent acquisition site. The blended model achieved an ROC AUC of ~0.66 using features extracted from anatomical data only. One observation from this effort was the fact that prediction accuracy increased with increasing sample size. Another was that while prediction accuracy for the subset of unseen data was similar to validation accuracy, accuracy for the novel site was poorer, illustrating the challenge of generalization, particularly to new data collection sites.

Although recent gains in prediction accuracy are promising, Machine Learning studies conducted to date have two main limitations. The first is that preprocessing pipelines often have many steps, each of which can introduce biases to prediction models. In particular, preprocessing typically includes transformation to a template space, such as MNI152, which was created using anatomical scans acquired from neurotypical adults. Template normalization may therefore negatively impact the ability to detect Autism-related alterations in brain structure, introduce biases, and lead to poorer reproducibility[30]. A second limitation is that datasets used for prediction tend to be clinically heterogeneous, but this heterogeneity is not explicitly accounted for in the models, leading to inconsistent results between separate datasets[40]. Many Autistic participants have a secondary diagnosis, which is often another psychological condition such as ADHD or anxiety, or a neurological condition such as epilepsy or Fragile X syndrome[17,24,26]. Ignoring these comorbidities may introduce biases or lead to non-specific biomarkers[17], since in such analyses, the label “autism” is not well delimited.

In the current study, we sought to develop a prediction pipeline that could overcome these challenges. To do this, we trained 3-Dimensional Deep Learning models to predict Autism diagnosis from minimally preprocessed structural MRI data, to avoid biases introduced by template normalization. To address the influence of clinical heterogeneity, we built our models using a large sample of 1329 patients (521 with autism) without comorbidities, following the classical framework of train-validate-test. To test if the patterns identified by the best models were robust to comorbidity, we tested the three best models on a second dataset comprising 270 patients (155 with autism) with comorbid diagnoses.

Deep Learning models can extract meaningful implicit features during the optimization process, which minimizes the preprocessing required and ultimately reduces prediction time. While 2D Deep Learning models are increasingly popular, 3D Deep Learning is not widely used in Medical Imaging applications, in part because of the large number of parameters to optimize (greater than in 2D) and concerns related to interpretability. To address the challenge of extracting information about predictive features (i.e., interpretability), we leveraged recently developed methods to build an interpretation pipeline that identifies predictive brain areas while avoiding the requirement for template normalization.

In this paper, we described our novel pipeline for interpretable 3D Deep Learning prediction of Autism diagnosis from structural MRI data. In our proof-of-concept analyses, our models achieved the same prediction accuracy as is typical for Machine Learning models, while avoiding the potential biases introduced by template normalization. Our interpretation pipeline identified a set of regions that replicated well across datasets (including participants with comorbidities), and models, and which converged with previous structural imaging studies on Autism. To facilitate further development of our pipeline, we have openly shared all our code through GitHub (https://github.com/garciaml/Autism-3D-CNN-brain-sMRI).

## 3 Materials and Methods

### 3.1. Data and Quality Control

We used T1-weighted structural MRI data from the ABIDE I (980 scans) and II (857 scans) datasets[31-32] and 140 scans from ADHD200[41]. We performed quality control using BrainQCNet[42], retaining scans with a probability score below 60% as advised in [42]; 797 scans from ABIDE I, 704 from ABIDE II and 98 from ADHD200 remained after this step.

Our primary analysis focused on participants with a diagnosis of Autism but no reported comorbidity and comparison participants with no psychiatric diagnosis. Excluding participants with comorbidities resulted in a dataset of 1329 participants which were used for training, validating and testing the models.

All participants in the testing set (*n* = 65, 26 with Autism) were obtained from different (independent) data collection sites than participants in the training (*n* = 1074, 421 with Autism) and validation (*n* = 190, 74 with Autism) sets.

To examine the impact of comorbidities on prediction accuracy, we created a second evaluation set of participants who had at least other diagnosis in addition to Autism, such as ADHD, phobias, depression, and anxiety. This dataset (testing set 2) contained scans from 270 participants (155 with Autism diagnosis).

Further details on the datasets are provided in **S1 - Detailed Data Description**.

### 3.2. Preprocessing

We employed a minimal preprocessing pipeline that did not apply transformation to template space, to avoid any impact of brain normalization on the detecting of Autism-related alterations in brain structure. Instead, we applied FSL’s Brain Extraction Tool (BET; https://fsl.fmrib.ox.ac.uk/fsl/fslwiki/BET) to remove non-brain tissue, followed by a number of minor non-deforming transformations, to prepare our data to be processed by the Deep Learning algorithm:

- *<u>Resolution homogenization</u>*: the ABIDE datasets comprise data from different data collection sites, each of which has different scanners and acquisition protocols, Accordingly, the T1-weighted volumes have heterogeneous voxel spacing that could bias the analysis. We used Linear Interpolation to perform resampling, with the Resample function from the Python library TorchIO (https://torchio.readthedocs.io/_modules/torchio/transforms/preprocessing/spatial/resample.html#Resample), built from the Insight Toolkit (https://itk.org/Doxygen/html/index.html) to resample all volumes to a fixed resolution of 1.5mm*1.5mm*1.5mm. We also reordered the data to RAS+ orientation.
- *<u>Intensity normalization</u>*: We removed the noise generated by voxel value outliers in every image by truncating the intensities to the range of 0.5 to 99.5 percentiles using the RescaleIntensity function from TorchIO. We also normalized each volume by subtracting the mean intensity value ν_*m*_ to each voxel value ν_*i*_, and then dividing by the standard deviation ν_*sd*_, obtaining a new voxel value ν′_*i*_.

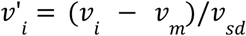
- *<u>Cropping or Padding</u>*: We cropped or padded each volume to obtain a uniform shape for all the volumes of 256*256*256. This shape was sufficiently large to fit the full brains and was also appropriate as an input shape to our deep learning models, in view of the filters applied all along each network (described in detail below).

### 3.3. Classification Models in 3D

Comparing different types of algorithm enables the detection of overfitting and retention of the best type of algorithm for the given problem[43]. We compared two models: (1) DenseNet121[44] and (2) Med3D-ResNet50[45], well known Deep Learning algorithms with good 2D performance[44-45]. DenseNet121 is more compact and has fewer parameters than ResNet50 making it possible to train on 3D data, while Med3D-ResNet50 [45] is a version of ResNet50 that has been pre-trained on medical images, including brain sMRI scans. Logically, pre-trained models enable better convergence and performance on new data and tasks of the same context. We fine-tuned Med3d-ResNet50 to adapt it to our task by training the last convolutional layers (corresponding to the 4th convolutional block). We also appended the last classifier block, consisting of a global average pooling layer and a fully connected layer (see **S2 - Model architectures**)

Like in [44] and in [45], we used the ReLU function as the activation function, the cross entropy loss, and the Adam optimizer with a fixed learning rate of 0.001.

### 3.4. Interpreting outcomes of Deep Learning algorithms

#### 3.4.1. Guided-Grad-CAM

In order to interpret and evaluate the reliability and relevance of our 3D Deep Learning models, we used Guided Grad-CAM[46], which combines guided backpropagation[47] and Grad-CAM[46]. This represents a good trade-off between the precision offered by feature maps produced by interpretability algorithms and the processing time required. Mathematically, guided Grad-CAM[46] is an element-wise product of the results of the two algorithms. It returns a high resolution map of the fine-grained features that is also class-discriminative.

In the context of our study, for a given trained CNN model (either DenseNet121 or Med3DNet-ResNet50), we used guided Grad-CAM to generate one “attention map” for each participant at the inference step (i.e. the first layer of the CNN). This attention map matched the input scan resolution and voxel dimensions, and its voxel values corresponded to scores of “importance” for the prediction of Autism/non-Autism by the trained CNN model. Mathematically, for a given input participant’s scan, we computed *q*_50%_ - the median of the voxel values of the attention map obtained with guided Grad-CAM. We then built a binary mask by returning all the voxel values lower than *q*_50%_ to 0 and all voxels greater than *q*_50%_ to 1. We used this mask *M* to identify the brain regions that are the most important for the prediction of Autism across the sample and across algorithms.

#### 3.4.2. HighRes3DNet

As noted above, a key feature of our preprocessing pipeline was our avoidance of normalization to a group template. This creates a significant challenge for the identification of the brain areas that were most predictive of diagnosis across participants. We solved this challenge by segmenting individual scans into anatomical units and combining this information with the mask *M* created in the preceding step.

HighRes3DNet[48] is a Deep Learning algorithm that segments brain MRI scans following the GIF brain parcellation (V3, http://niftyweb.cs.ucl.ac.uk/program.php?p=GIF ; [49]). The GIF algorithm was especially built to be robust to brain morphological differences, especially those encountered in populations with atypical brain development like Autism[49].

We segmented each participant’s brain with the HighRes3dNet algorithm (first homogenizing scans to voxel size 1mm*1mm*1mm using linear Interpolation). The resulting segmented images were resampled to 256*256*256 images of voxel size 1.5mm*1.5mm*1.5mm to match the resolution of the attention maps obtained from the guided Grad-CAM algorithm, while retaining the segmented voxel values.

Specifically, we know that the information on the transformations applied to the segmented image is contained into the affine matrix of the resulting transformed segmented image.

Mathematically, we note *X* = [*x, y, z*, 1], the column vector of the coordinates x, y, z of a voxel in a segmented image obtained with HighRes3DNet (voxel size: 1mm*1mm*1mm), *Y* = [*x*′, *y*′, *z*′, 1] the column vector of the coordinates x’, y’, z’ of a voxel in the corresponding transformed segmented image (size: 256*256*256; voxel size: 1.5mm*1.5mm*1.5mm), and *A* ∈ |*R*^4^ its affine matrix. We note *B*, the inverse matrix of *A*, such that *BA* = *A*^−1^*A* = *I*, where *I* is the identity matrix in *R*^4^.

Thus, we have the relationship:

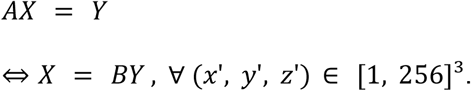

Thus, if we take *x*′, *y*′, *z*′ the coordinates of a voxel in the mask *M* obtained from guided Grad-CAM, we can obtain the corresponding *x, y, z* voxel coordinates in the segmented image, and thus get the voxel value and the name of the area at (*x, y, z*).

Applying this procedure for every scan, we obtained a table containing, for every area of the HighRes3DNet atlas, a relative frequency corresponding to the number of voxels in the area with value = 1, divided by the total number of voxels in this area in the segmented image. This relative frequency corresponds to the proportion of the area that is considered important for the prediction by a CNN model, for that participant. These proportions were then used to compare different brain areas and to draw up a ranking of brain areas for each model, dataset (training, validation, testing sets), and type of prediction (True Positives, True Negatives, False Positives, False Negatives), to improve interpretability for our CNN models.

### 3.5. Machine and Code availability

We trained our model on a GPU Nvidia RTX 3090 (24 GB memory) with a batch size of 2. We openly shared the code of this project on GitHub, in the repository: https://github.com/garciaml/Autism-3D-CNN-brain-sMRI. The models are also shared so that they can be reused as pre-trained models for similar applications.

## 4. Results

### 4.1. Training performance

For all the probability scores of all the models, we chose a threshold of 0.5 for the class “Autism diagnosis” to define the prediction and compute the accuracy and ROC AUC scores. We trained each model up to 100 epochs and computed model accuracy using the validation set (190 scans) every two epochs. Details on the validation set accuracy during training for the two models DenseNet161 and Med3d-ResNet50 are provided in **S3 Fig.1** in **S3 - Performance of the models**.

For ResNet50, the best validation set accuracy was 62.6%, achieved at 42 epochs. For DenseNet121, 66.3% accuracy was achieved at 32 epochs and 67.4% was achieved at 70 epochs. Next, we compared the performance of these three best models (one ResNet50 model and two DenseNet121 models) for the prediction of diagnosis in the training, validation, and testing sets.

### 4.2. Prediction Performance: Autism diagnosis

For the prediction of Autism diagnosis, the three best models behaved differently, as shown by the Receiver Operating Characteristic curves in **Fig 1**. Med3d-ResNet50-42ep overfitted the data - the accuracy and ROC AUC scores were very high on the training set (94.2% and 99.9% respectively) but much lower on the validation (acc = 62.6% and AUC = 62.1%) and testing sets (acc = 53.8% and AUC=57.3%). DenseNet121-32ep appeared to be more stable in terms of its overall performance on the training (acc = 65.5% and AUC = 69.1%), validation (acc =66.3% and AUC = 68.8%) and testing (acc =55.4% and AUC = 60.7%) sets. DenseNet121-70ep had better performance on the training (acc = 69.7% and AUC = 77.1%) and validation (acc = 67.4% and AUC = 68.1%) sets than DenseNet121-32ep, but poorer performance on the testing set (acc = 40% and AUC = 38.1%).

**Fig 1.**
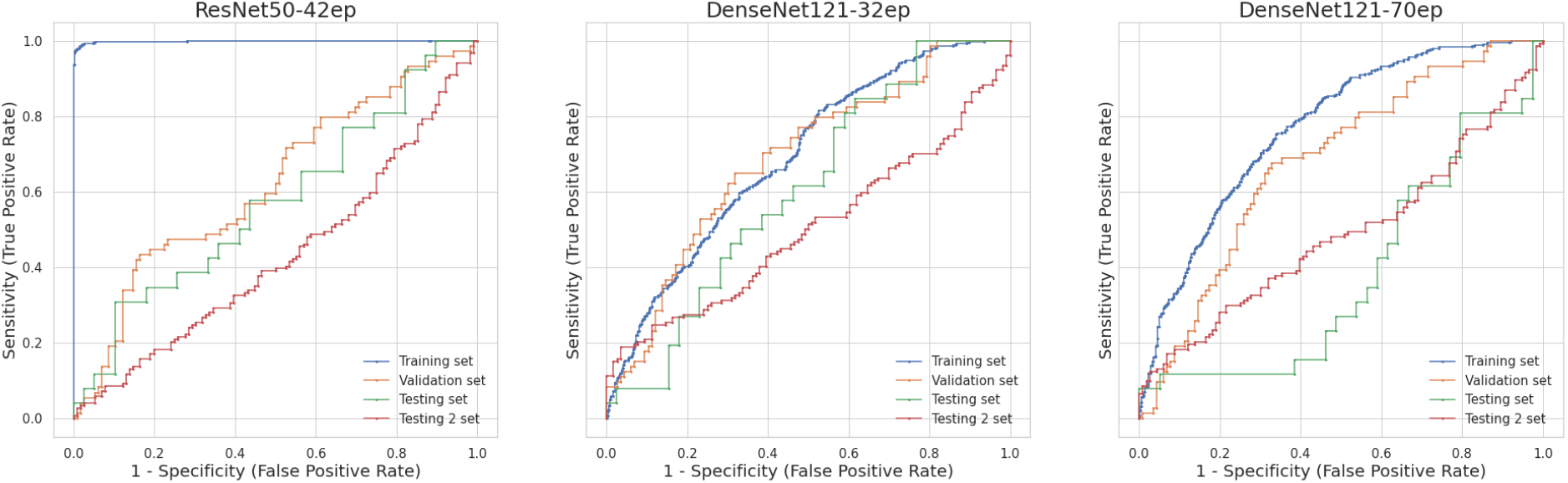
Receiver Operating Characteristic curves for all the three models and all the four datasets

Table 1 displays the sensitivity and specificity of each model for each dataset. DenseNet121-32ep exhibited high specificity on the training and validation sets, but low sensitivity. Paradoxically, it had high sensitivity but low specificity on the testing set. DenseNet121-70ep behaved similarly on the testing set while on the training and validation sets, sensitivity and specificity were balanced and fairly high. Finally, for Med3d-ResNet50-42ep, sensitivity and specificity were very high on the training set, unbalanced on the validation set with low sensitivity and very high specificity, and balanced on the testing set, but with moderate values.

**Table 1.** Sensitivity and Specificity of each model on each dataset (training, validation, testing sets with no comorbidity and testing set 2, which included patients with comorbidities).

|  | Med3dNet - Resnet50;<br>trained on 42 epochs | DenseNet121;<br>trained on 32 epochs | DenseNet121;<br>trained on 70 epochs |
| --- | --- | --- | --- |
| Sensitivity | Train: 85,3%<br>Validation: 17,6%<br>Test: 50%<br>Test 2: 8,4% | Train: 32,8%<br>Validation: 36,5%<br>Test: 84,6%<br>Test 2: 7,6% | Train: 68,2%<br>Validation: 66,2%<br>Test: 69,2%<br>Test 2: 31% |
| Specificity | Train: 100%<br>Validation: 91,4%<br>Test: 56,4%<br>Test 2: 87,8% | Train: 86,7%<br>Validation: 85,3%<br>Test: 35,9%<br>Test 2: 100% | Train: 70,8%<br>Validation: 68,1%<br>Test: 20,5%<br>Test 2: 73% |

Sensitivity on the second testing set, which included participants with comorbidities, was low for all models. This demonstrates that when the training and testing sets include only participants without known comorbidities, predicting Autism diagnosis for participants with comorbidities is particularly challenging. Here, we found that this produces a large increase in False Negatives in particular. One potential explanation is that neuroimaging markers of Autism are less salient when individuals have another diagnosis involving similar or other neuroimaging markers. Another explanation is that more data is needed to adequately train DL algorithms on the whole spectrum of Autism in the context of comorbidities.

Further details and comments on the performance of the models are given in **S3 - Performance of the models**, and a comparison of the predicted scores with the scores of diagnosis are given in **S4 - Analysis of ADI-R and ADOS scores, age, gender and full IQ**.

### 4.3. Interpretability: *True Positive* discriminative ROIs

We segmented each participant’s scan using HighRes3DNet (GIF parcellation), to extract a measure of “prediction importance” (the output of the guided Grad-CAM algorithm) for each of the three best models. We identified the regions that best contributed to True Positives (TP), True Negatives (TN), False Positives (FP) and False Negatives (FN), across the whole dataset (training + validation + testing 1 & 2 sets).

For every pair of model and dataset, we defined the “most predictive” regions as those with relative frequency values (see **Section 3.6**, above) greater than the 90% percentile. This yielded 16 regions for each model and dataset pair. To compare the most predictive regions across models and datasets (training, validation, testing Set 1 - no comorbidities, testing set 2 - with comorbidities), we summed the presence (1) or absence (0) of the most predictive regions over all the datasets, separately for *True Positives* and *True Negatives*. Across all three models, 79 areas were found to be most predictive for *True Positives*, including 26 areas spanning both left and right hemisphere, 23 areas in the left hemisphere only, 3 areas in the right hemisphere only, and the Corpus Callosum. Retaining only areas that replicated across all four datasets (training, validation, and test 1/2), we found that areas in the left hemisphere were more replicable than those in the right, and that the majority of areas were in the prefrontal cortex. The **S5 - Most important regions for the prediction of True Positives** provides **S5 Table 6** that summarizes the most replicable regions across models and datasets that are important to predict *True Positives*, and a detailed analysis of these most replicable regions.

Overall, 17 regions were found to best predict *True Positives* across models and replicate across datasets (training, validation, and testing 1/2). These regions are shown in **Fig 2** and include regions in the left frontal lobe (medial frontal cortex, inferior and middle frontal gyrus, lateral and medial precentral gyrus, anterior and subcallosal cingulate gyrus, and posterior orbital gyrus), left temporal lobe (temporal pole, planum temporale, parahippocampal gyrus), parietal lobe (parietal operculum, supramarginal gyrus, and superior parietal lobe), as well as left parietal white matter and the right ventral thalamus.,

Looking at these data another way, and taking the regions that were most predictive across datasets and which replicated across the three models, we again obtained left hemisphere regions that are located in the frontal lobe - middle and inferior frontal gyrus (pars triangularis) and medial precentral gyrus - and in the limbic system and its associated structures - anterior cingulate gyrus, subgenual cingulate gyrus, parahippocampal gyrus (**Fig 2b**).

**Fig 2.**
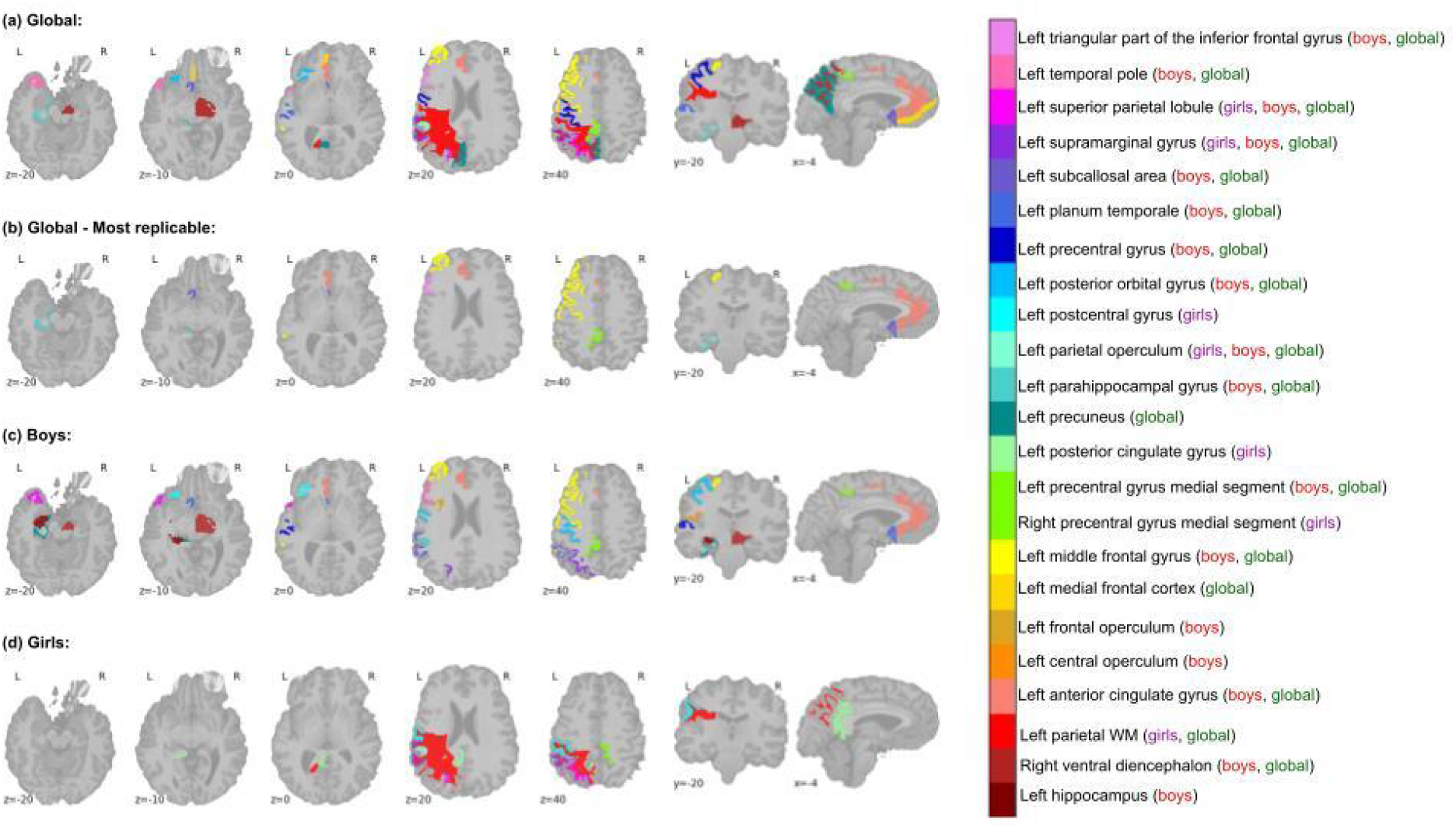
(a) Regions most predictive of Autism diagnosis; (b) Most predictive regions that replicate across datasets, (c) Most predictive regions for boys; (d) Most predictive regions for girls.

#### 4.3.1. Effect of gender

Regions important for predicting *True Positives* for boys were different from those for girls. Regions common to both genders were located in the left parietal lobe: parietal operculum, supramarginal gyrus, and superior parietal lobule (**Fig 2c, d**). Globally, regions found important to predict *True Positives* for boys were more replicable across the datasets (training, validation, testing 1/2) than for girls. For boys, several left prefrontal regions were replicably predictive of Autism diagnosis: left anterior cingulate gyrus, middle frontal gyrus, inferior frontal gyrus (pars triangularis; ResNet50-42ep only), medial precentral gyrus (DenseNet121-32ep) and precentral and parahippocampal gyrus (DenseNet121-70ep).

**S8 - True Positives by Gender** shows these results in **S8 Tables 10 and 11**.

### 4.3.2 Relationship with age

Autism has been associated with disrupted brain development across the lifespan. To assess whether there were any developmental trends in the most predictive areas, we created four age categories (5-10yrs, 10-15yrs, 15-20yrs, >20yrs) and identified the most predictive (True Positives) regions for each category, separately for boys and girls. **S 9 - True Positives by Gender and Age** shows these results.

Our results showed that the most discriminative regions varied with the age. In particular, left precentral gyrus, central operculum, and posterior orbital gyrus replicably predicted *True Positives* in boys aged 5-10yrs, while left inferior frontal gyrus (pars triangularis), subcallosal/subgenual cingulate cortex, and supramarginal gyrus, were most predictive for boys aged 10-15 years old.

In addition, we found that the replicability of each region decreased as age increased. Indeed, we found that the left and inferior frontal (pars triangularis) gyrus, posterior orbital gyrus and putamen were most predictive for 15-20 years old, but only for participants without comorbidities. Left temporal areas - parahippocampal gyrus, superior temporal gyrus and temporal pole - were most predictive for males aged 20-64yrs without comorbidity.

Examining global prediction performance for these different age groups reveals other interesting trends, such as a decrease in the number of False Negatives and True Negatives with increasing age, for both boys and girls. This suggests that our prediction of Autism diagnosis tended to be more sensitive but less specific as age increased.

#### 4.3.3. True Negatives

We adopted the same approach described above to identify regions most predictive of *True Negatives* (i.e., absence of an Autism diagnosis). The results (see **S6 - Most important regions for the prediction of True Negatives**) showed that the most replicable regions for predicting *True Negatives* were in the left hemisphere and included the frontal operculum, the precuneus, the planum polare, the inferior occipital gyrus, the occipital fusiform gyrus, the superior occipital gyrus and the thalamus proper. It also included the cerebellar vermal lobules VI and VII.

Another result is that the regions left precuneus, parietal operculum, and superior parietal lobe, and right thalamus were important (at various degrees of replicability and for different models) for the prediction of both *True Negatives* and *True Positives*. The 23 other regions important for the prediction of True Negatives are different from those that were important to the prediction of True Positives.

#### 4.3.4. Bad predictions - False Positives and False Negatives

We adopted the same approach described above to identify regions most predictive of *False Positives* (i.e., incorrectly predicted Autism diagnosis) and *False Negatives* (i.e., incorrectly failed to predict Autism diagnosis). **S7-Most replicable regions for *False Positives* and *False Negatives*** shows these results. No highly replicable regions (replicable over all datasets) were found for *False Positives*. However, regions with a high level of replicability for *False Positives* for DenseNet121-70 overlapped with replicable regions for the prediction of *True Positives* for the two other models and included the middle frontal gyrus, precentral gyrus medial segment, and triangular part of inferior frontal gyrus. This illustrates differences in the calibration of each algorithm and demonstrates the importance of comparing different models. For *False Negatives*, the most replicable regions were again found in the left hemisphere and included the left frontal operculum, left precuneus, left superior temporal gyrus, left planum polare, left inferior occipital gyrus and left occipital fusiform gyrus.

### 4.4. Does image background contribute to model predictions?

As a final test, we examined whether image background (i.e., information outside the brain) contributed to predictions. For Med3d-ResNet50-42ep the relative frequency of the Background (RF) is the smallest (RF=0.97%) and the second smallest for DenseNet121-70ep (RF=0.28%), meaning that this area is not considered predictive for the models. For DenseNet121-32ep, it is among the last 4% informative areas of the model (RF=0.74%). These results confirm that the models use information from inside rather than outside the brain to make a prediction, supporting their validity.

### 4.5. Multi-site effect

We observed an inhomogeneous consistency of the distributions of probability scores between the different sites (see **S10 - Multi-site effect**). We displayed the accuracy scores for every site in the whole dataset (training+validation+testing sets) in **S10 Table 20**, and it also confirmed the multi-site effect.

## 5. Discussion

This study outlines and demonstrates a novel approach for inferring Autism diagnosis from structural brain imaging data using 3D Deep Learning algorithms. To maximize the interpretability of the model outputs, we also used a second type of algorithm - guided Grad-CAM[46] - to extract patterns important for the predictions. This step revealed a set of regions predominantly located in the left hemisphere, including lateral and medial prefrontal cortex, anterior cingulate, the superior temporal gyrus, lateral parietal regions including supramarginal gyrus, parahippocampal gyrus. The only right hemisphere region highlighted in our analyses was the right thalamus. The regions highlighted by this interpretability analysis, the brain structural features of which were most important for accurate inference of Autism diagnosis (i.e., *True Positives*), are highly consistent with the literature. Our predictive modeling framework has considerable potential to be extended to further datasets to identify and refine sensitive and specific brain biomarkers of Autism using MRI data.

### 5.1. 3D Deep Learning applied to minimally processed data

To our knowledge, this is the first time that 3D-DL CNNs have been used to predict Autism diagnosis from 3D structural MRI scans. Our findings show that these algorithms are capable of inferring Autism diagnosis on the basis of structural MRIs with at least the same level of accuracy as traditional Machine Learning algorithms, while requiring a smaller number of training epochs. The average accuracy score (64.1%) and ROC AUC score (0.67) obtained for participants without comorbidities is consistent with previous Machine Learning models trained on sMRI data (e.g., [39]). The comparable accuracy we achieved should be viewed in the context of the speed of inference of Deep Learning models over Machine Learning approaches. While Machine Learning algorithms require inputs derived following extensive preprocessing of structural MRI data, including normalization to template space, our Deep Learning models used minimally preprocessed data. In particular, we avoided transformation to template space, a near-universal requirement of neuroimaging analyses that may negatively impact the ability to detect structural alterations associated with the diagnosis of interest. Although our pipeline included some minimal preprocessing steps to address the fact that a diversity of scanners and acquisition protocols was used across data collection sites, resulting in heterogeneous voxel spacing and signal intensities. Resolution homogenization and intensity normalization were applied to address these variations, and it is possible that these steps could bias the algorithm. Further, despite these steps, a clear effect of the data collection site was observable. Future studies will incorporate specific preprocessing steps like the ComBat algorithm[50] to integrate scan parameters during training and minimize site effects.

### 5.2. Interpretability

The outputs of Deep Learning models are not straightforwardly understandable, giving rise to the challenge of poor interpretability. This challenge arises because mathematically, Deep Learning models are composed of multiple functions. Each of these functions is nonlinear and is itself the sum of multiple functions. Further, models such as the 3D CNNs used in the current study have a large number of parameters that must be optimized. One of the goals of our study was to address this drawback by devising a pipeline that would allow for the extraction of predictive brain regions, providing interpretability. Guided Grad-CAM[46] was chosen for this purpose, due to its reasonable computation time and its ability to return fine-grained class-specific segmentations of important (predictive) voxels in the input images.

A challenge for our novel interpretability process was to identify brain areas that were predictive of Autism diagnosis across participants while avoiding the requirement for template normalization. To address this issue, we used a segmentation algorithm to partition individual volumes into established anatomical regions. We used HighRes3DNet[48] for this task because it was built to be pathology-agnostic, robust to brain morphology differences, and has reduced computation time compared to other algorithms (e.g., the GIF algorithm[49]). We performed a detailed analysis of the regions that were most relevant for inferring an Autism diagnosis, by examining true and false positives and negatives separately for each dataset and algorithm. We also identified regions that were reproducibly identified across algorithms and datasets. This detailed analysis is important because each model has biases, likely resulting in a differential weighting of anatomical features and brain areas. This analysis showed that regions of left prefrontal cortex (inferior and middle frontal gyrus, medial prefrontal gyrus, anterior and subgenual cingulate cortex), along with the parahippocampal gyrus were brain regions whose morphological features contributed most to the accurate inference of Autism across models and datasets without and with comorbidities. The areas highlighted are consistent with previous studies reporting Autism-related disruptions to cortical development[24, 51-53] and gyrification processes[24, 54] in these regions. Further, also consistent with the literature, we found that the most predictive regions varied according to both gender and age, as well as the presence of comorbidities[17, 24, 55]. This is consistent with observations that Autism is a complex condition, with patterns of neurological divergence that vary with age[17, 24, 52-53] and sex[17, 24, 55].

Reproducibly predictive regions in the limbic system (left parahippocampal gyrus, anterior cingulate gyrus, and subcallosal area), dorsal medial frontal cortex, and precentral gyrus fit well with previous work on the role of atypical socio-emotional and motor circuitry in Autism[56-60]. Many of the left-hemisphere regions identified as contributing to accurate inference of Autism diagnosis fall within the canonical left-lateralized language network, including inferior prefrontal and inferior parietal regions, and the planum temporale in superior temporal gyrus[61-63]. Divergent structure and function in the language network is a robust and reproducible finding in Autism[64-67]. Since early language processing appears to be an important predictor of long-term outcomes in Autism[68-70] identification of early-emerging structural alterations in the underlying language network has the potential to yield a powerful marker of Autism or Autism subtypes, which could, in turn, direct individualized interventions and improve prognosis.

An important caveat is that while our novel interpretation step identified which regions of the brain had morphological features relevant to the model-based inference of Autism, it did not provide information on what these morphological features were. For example, features such as cortical thickness, the location of the gray-white boundary, surface area, and gyral/sulcal morphometry could all play a role in prediction of Autism[22,52,71]; and different morphological features may be relevant in different brain areas. While the precise nature of the Autism-related morphological features are not discernable from our analyses, our predictive modeling analyses can be followed up with in-depth, targeted, and hypothesis-driven examinations of the areas highlighted in independent samples to uncover the nature of these features.

### 5.3. Limitations

Our pipeline for prediction of neuropsychiatric diagnosis (Autism) on the basis of minimally preprocessed T1 MRI scans advances progress toward interpretable 3D Deep Learning applications in biological psychiatry and toward the identification of reproducible brain biomarkers that will help refine diagnoses and treatment plans across conditions. Our study had several limitations, however, which may be addressed in further refinements of our pipeline.

First, we trained our models on 100 epochs, which is an acceptable number relative to other studies using 3D MRI scans[72], but which may have limited the convergence and optimization of the algorithms. Future work may train on a larger number of epochs or may employ earlystopping[73] to optimize training. Using the entire structural MRI scans (to explore prediction across the whole brain) may also have posed a challenge for convergence towards the “True” solution. Further, although we used a large dataset (1074 participants to train the models, 525 to validate and test the models), the amount of data available is still rather limited when we consider the clinical heterogeneity of Autism. This idea is supported by the poor prediction performance we observed for test set 2, which included participants with comorbid diagnoses (average accuracy = 46.3%, ROC AUC = 0.47 and average sensitivity = 15.7%). There are still questions in the literature about whether predicting a binary label, “Autism vs non-Autism” is a useful or appropriate endeavor, since Autism is a wide spectrum of behaviors and abilities which may encompass as many as four subtypes[23], and there is also considerable overlap of symptoms and neuromarkers across psychological conditions[17]. Future analyses will need to leverage even larger datasets to better address the clinical heterogeneity of Autism and to explore the prediction of categories beyond Autism and non-Autism.

Another limitation is related to the segmentation algorithm we used in the interpretation step. We used HighRes3DNet[48] to obtain rapid segmentation for each brain using the GIF algorithm[49], which was built to be robust on atypically developing brains. The segmentation produced is rather coarse, however - the algorithm outputs relatively large parcels, encompassing anatomically heterogeneous regions such as the anterior cingulate gyrus or superior parietal lobule. Further, as noted above, while our interpretation process localized regions that were important for prediction of Autism, it did not provide information on what the predictive morphological features of those regions were.

### 5.4. Future Directions

There is considerable scope to extend our interpretable Deep Learning pipeline to the prediction of other neurological or neuropsychiatric conditions or to other MRI modalities. Traut et al.[39] reported that prediction of Autism was considerably improved (from AUC=0.66 using only anatomical MRI to AUC=0.79 using both anatomical and functional data) for a blended model that incorporated both functional and structural MRI data. Future work will examine whether functional MRI data can also improve our models. Other efforts to improve our model will include training the models on more epochs, exploring other architectures, integrating scanning parameters and other confounds such as gender and age, and using different and extended class labeling. We have shared all our code (https://github.com/garciaml/Autism-3D-CNN-brain-sMRI) to enable other researchers to apply, reuse, and further develop our models and approach.

## 6. Conclusion

In this paper, we described a novel methodology to build a predictive model to infer Autism diagnosis using 3D Deep Learning applied to structural MRI scans, coupled with an interpretation step in the form of a descriptive method that identified the brain regions that were most important for accurate inference. Importantly, we applied our models to minimally preprocessed data - completely avoiding the template normalization step, which may obscure diagnosis-related alterations in brain structure. We found that the predictive performance of our models was equivalent to that of Machine Learning models reported in the literature, while requiring less time to generate predictions (due to minimal preprocessing). There is considerable scope to refine our method or to incorporate other modalities (e.g., fMRI) to further boost predictive performance.

Our method for interpreting the output of Deep Learning models revealed highly predictive brain regions that were consistent with the literature, demonstrating that 3D Deep Learning models produce biologically plausible results without a priori knowledge or the requirement for pre-computation of morphological derivatives (e.g., volumes, cortical thickness, surface area). Although challenges related to the clinical heterogeneity of Autism remain to be addressed, we have openly shared our code and models for others to build on and extend, and to further progress the field towards the identification of robust and reproducible brain biomarkers for neuropsychiatric conditions.

## Data Availability

All data produced in the present work are contained in the manuscript.

https://github.com/garciaml/Autism-3D-CNN-brain-sMRI

## Abbreviations

CNN: Convolutional Neural Networks, a category of Deep Learning algorithm
ML: Machine Learning
DL: Deep Learning
Med3dNet - Resnet50: pretrained Residual Networks model with 50 layers
DenseNet121: Densely Connected Convolutional Networks with 121 layers
Epoch: a hyperparameter that defines the number of times that the learning algorithm has optimized the parameters on the entire training dataset.
MRI: Magnetic Resonance Imaging

## 7. Acknowledgements

We have much gratitude and appreciation for the Irish Research Council and the Hypercube Institute (Paris) for their funding.

We also thank Pr. Louise Gallagher as well as Dr. Robert Whelan for their feedback and guidance.

## 8. Disclosure of competing interests

None.

## Notes

### Competing Interest Statement

The authors have declared no competing interest.

### Funding Statement

This study was funded by an Irish Research Council Postgraduate Scholarship and a donation for this research project from a French non-profit
endowment fund called the HyperCube Institute (IHC). The IHC had to terminate all
activity in 2020 due to the impacts of COVID-19.
The funders had no role in study design, data collection and analysis, decision to
publish, or preparation of the manuscript.

### Author Declarations

The databases used in the project - ABIDE 1 and 2, ADHD200 - are shared by the International Neuroimaging Data-sharing Initiative which established rules priorly of making every dataset publicly available. Each dataset had to be fully de-identified and anonymized data in accordance with the U. S. Health Insurance Portability and Accountability Act (HIPAA). All the datasets have been collected following the local region regulations on ethics and data protection. Each research group tailored specific agreements on data reuse for their participants. Data usage is unrestricted for non- commercial research purposes, it is openly shared with the scientific community under the license Creative Commons BY-NC-SA. My work with these open data is approved by the School of Psychology Research Ethics Committee at Trinity College Dublin.

