## Supporting information for "Towards 3D Deep Learning for neuropsychiatry: predicting Autism diagnosis using an interpretable Deep Learning pipeline applied to minimally processed structural MRI data"

### S1 - Detailed data description

| | | No comorbidity | | | At least one comorbidity |
|---|---|---|---|---|---|
| Dataset | Data-collecting site | **Train set (1074 sub.)** | **Validation set (190 sub.)** | **Testing set (65 sub.)** | **Testing set 2 (270 sub.)** |
| **ABIDE I** | CALTECH | Total: 30 (Autism: 16; No Autism: 14) | Total: 6 (Autism: 2; No Autism: 4) | | |
| | CMU | Total: 25 (Autism: 13; No Autism: 12) | Total: 2 (Autism: 1; No Autism: 1) | | |
| | KKI | Total: 22 (Autism: 6; No Autism: 16) | Total: 3 (Autism: 0; No Autism: 3) | | Total: 9 (Autism: 7; No Autism: 2) |
| | LEUVEN_1 | | | Total: 27 (Autism: 14 ; No Autism: 13) | |
| | LEUVEN_2 | Total: 28 (Autism: 12; No Autism: 16) | Total: 3 (Autism: 1; No Autism: 2) | | |
| | MAX_MUN | Total: 30 (Autism: 10; No Autism: 20) | Total: 3 (Autism: 3; No Autism: 0) | | |
| | NYU | Total: 103 (Autism: 27 ; No Autism: 76) | Total: 21 (Autism: 5; No Autism: 16) | | Total: 28 (Autism: 28; No Autism: 0) |
| | OHSU | Total: 19 (Autism: 10; No Autism: 9) | Total: 3 (Autism: 1; No Autism: 2) | | |
| | OLIN | Total: 12 (Autism: 8; No Autism: 4) | Total: 7 (Autism: 4; No Autism: 3) | | |
| | PITT | Total: 31 (Autism: 15; No Autism: 16) | Total: 8 (Autism: 4; No Autism: 4) | | |
| | SBL | Total: 26 (Autism: 13; No Autism: 13) | Total: 3 (Autism: 1; No Autism: 2) | | |
| | SDSU | Total: 10 (Autism: 1; No Autism: 9) | Total: 2 (Autism: 1; No Autism: 1) | | |
| | STANFORD | Total: 5 (Autism: 1; No Autism: 4) | Total: 2 (Autism: 2; No Autism: 0) | | |
| | TRINITY | Total: 36 (Autism: 18; No Autism: 18) | Total: 6 (Autism: 2; No Autism: 4) | | |

| | | | | | |
|---|---|---|---|---|---|
| | UCLA_1 | Total: 45 (Autism: 27; No Autism: 18) | Total: 7 (Autism: 5; No Autism: 2) | | |
| | UCLA_2 | Total: 13 (Autism: 6; No Autism: 7) | Total: 2 (Autism: 0; No Autism: 2) | | |
| | UM_1 | Total: 59 (Autism: 21; No Autism: 38) | Total: 13 (Autism: 6; No Autism: 7) | | |
| | UM_2 | Total: 28 (Autism: 12; No Autism: 16) | Total: 3 (Autism: 0; No Autism: 3) | | |
| | USM | Total: 54 (Autism: 35; No Autism: 19) | Total: 12 (Autism: 8; No Autism: 4) | | |
| | YALE | Total: 46 (Autism: 22; No Autism: 24) | Total: 4 (Autism: 2; No Autism: 2) | | |
| **ABIDE II** | BNI | Total: 8 (Autism: 7; No Autism: 1) | Total: 1 (Autism: 0; No Autism: 1) | | |
| | EMC | | | Total: 18 (Autism: 4; No Autism: 14) | Total: 9 (Autism: 9; No Autism: 0) |
| | ETH | Total: 25 (Autism: 7; No Autism: 18) | Total: 5 (Autism: 1; No Autism: 4) | | |
| | GU | Total: 56 (Autism: 19; No Autism: 37) | Total: 9 (Autism: 2; No Autism: 7) | | |
| | IP | Total: 29 (Autism: 7; No Autism: 22) | Total: 7 (Autism: 4; No Autism: 3) | | Total: 8 (Autism: 5; No Autism: 3) |
| | IU | Total: 31 (Autism: 15; No Autism: 16) | Total: 2 (Autism: 1; No Autism: 1) | | |
| | KKI | Total: 103 (Autism: 1; No Autism: 102) | Total: 20 (Autism: 0; No Autism: 20) | | Total: 36 (Autism: 32; No Autism: 4) |
| | KUL | Total: 12 (Autism: 12; No Autism: 0) | Total: 8 (Autism: 8; No Autism: 0) | | Total: 5 (Autism: 5; No Autism: 0) |
| | NYU 1 | Total: 36 (Autism: 12; No Autism: 24) | Total: 5 (Autism: 2; No Autism: 3) | | Total: 22 (Autism: 22; No Autism: 0) |
| | NYU 2 | Total: 5 (Autism: 5; No Autism: 0) | Total: 1 (Autism: 1; No Autism: 0) | | Total: 15 (Autism: 15; No Autism: 0) |
| | OHSU | Total: 47 (Autism: 11; No Autism: 36) | Total: 8 (Autism: 0 ; No Autism: 8) | | Total: 34 (Autism: 24; No Autism: 10) |

| | | | | | |
|---|---|---|---|---|---|
| | SDSU | Total: 51 (Autism: 28; No Autism: 23) | Total: 4 (Autism: 3; No Autism: 1) | | |
| | TCD | Total: 29 (Autism: 13; No Autism: 16) | Total: 7 (Autism: 3; No Autism: 4) | | |
| | UCD | | | Total: 20 (Autism: 8; No Autism: 12) | Total: 5 (Autism: 5; No Autism: 0) |
| | USM | Total: 20 (Autism: 11; No Autism: 9) | Total: 3 (Autism: 1; No Autism: 2) | | |
| **ADHD200** | Peking | | | | Total: 23 (Autism: 0; No Autism: 23) |
| | KKI | | | | Total: 10 (Autism: 0; No Autism: 10) |
| | NeuroIMAGE | | | | Total: 22 (Autism: 0; No Autism: 22) |
| | NYU | | | | Total: 65 (Autism: 2; No Autism: 63) |
| | OHSU | | | | Total: 20 (Autism: 0; No Autism: 20) |

**S1 Table 1.** Partition of ABIDE I, ABIDE II, and ADHD200 into training, validation and testing sets.

| | **Gender** | **Age** | **FIQ** |
|---|---|---|---|
| **Train** | Males: 853 | mean 17.159562<br>std 8.656338<br>min 5.128000<br>25% 11.005000<br>50% 14.653000<br>75% 20.100000<br>max 64.000000 | mean 110.290806<br>std 14.888248<br>min 41.000000<br>25% 101.000000<br>50% 111.000000<br>75% 121.000000<br>max 149.000000 |
| | Females: 221 | mean 15.026466<br>std 8.035651<br>min 5.220000<br>25% 9.789041<br>50% 12.361644<br>75% 16.800000<br>max 54.000000 | mean 111.308458<br>std 14.835831<br>min 66.000000<br>25% 101.000000<br>50% 113.000000<br>75% 122.000000<br>max 146.500000 |
| **Validation** | Males: 153 | mean 17.012265<br>std 8.623991<br>min 7.150000<br>25% 11.262800<br>50% 14.800000<br>75% 20.166667 | mean 110.043750<br>std 15.436532<br>min 49.000000<br>25% 100.750000<br>50% 112.000000<br>75% 119.250000 |

| | | | |
|---|---|---|---|
| | | max 64.000000 | max 147.500000 |
| | Females: 37 | mean 13.046654<br>std 5.848126<br>min 5.907000<br>25% 9.665753<br>50% 10.780822<br>75% 14.060000<br>max 32.000000 | mean 113.972222<br>std 14.624317<br>min 84.000000<br>25% 105.750000<br>50% 115.000000<br>75% 123.000000<br>max 149.000000 |
| **Test 1 (no comorbidity)** | Males: 57 | mean 17.087350<br>std 6.428793<br>min 7.129363<br>25% 10.663929<br>50% 17.416667<br>75% 22.000000<br>max 32.000000 | mean 109.976190<br>std 12.994348<br>min 83.000000<br>25% 101.500000<br>50% 108.500000<br>75% 118.250000<br>max 146.000000 |
| | Females: 8 | mean 12.005540<br>std 4.022715<br>min 6.395619<br>25% 8.400411<br>50% 13.500000<br>75% 14.520833<br>max 16.500000 | mean 113.200000<br>std 14.411801<br>min 92.000000<br>25% 105.000000<br>50% 120.000000<br>75% 122.000000<br>max 127.000000 |
| **Test 2 (with comorbidities)** | Males: 205 | mean 11.946206<br>std 5.006252<br>min 5.598000<br>25% 8.646575<br>50% 10.870000<br>75% 13.200000<br>max 35.000000 | mean 107.235632<br>std 15.966971<br>min 69.000000<br>25% 97.250000<br>50% 108.000000<br>75% 116.000000<br>max 148.000000 |
| | Females: 65 | mean 11.973690<br>std 5.715843<br>min 5.819000<br>25% 9.000000<br>50% 10.260000<br>75% 12.580000<br>max 38.760000 | mean 107.087719<br>std 12.884488<br>min 74.000000<br>25% 98.000000<br>50% 109.000000<br>75% 115.000000<br>max 132.000000 |

**S1 Table 2.** Gender breakdown and distribution of age and FIQ score for each dataset (training, validation, testing, testing 2 sets).

### S2 - Model architectures

| Layers | Output Size | DenseNet121 |
|---|---|---|
| Convolution | 128 x 128 x 128 | 7 x 7 x 7 conv, stride 2 |
| Pooling | 64 x 64 x 64 | 3 x 3 x 3 pool, stride 2 |
| DenseBlock 1 | 64 x 64 x 64 | 1 x 1 x 1 conv → 3 x 3 x 3 conv<br>x 6 |
| Transition Layer 1 | 64 x 64 x 64 | 1 x 1 x 1 conv |
| | 32 x 32 x 32 | 2 x 2 x 2 average pool, stride 2 |
| DenseBlock 2 | 32 x 32 x 32 | 1 x 1 x 1 conv → 3 x 3 x 3 conv<br>x 12 |
| Transition Layer 2 | 32 x 32 x 32 | 1 x 1 x 1 conv |
| | 16 x 16 x 16 | 2 x 2 x 2 average pool, stride 2 |
| DenseBlock 3 | 16 x 16 x 16 | 1 x 1 x 1 conv → 3 x 3 x 3 conv<br>x 24 |
| Transition Layer 3 | 16 x 16 x 16 | 1 x 1 x 1 conv |
| | 8 x 8 x 8 | 2 x 2 x 2 average pool, stride 2 |
| DenseBlock 4 | 8 x 8 x 8 | 1 x 1 x 1 conv → 3 x 3 x 3 conv<br>x 16 |
| Classification Layer | 1 x 1 x 1 | 8 x 8 x 8 global average pool |
| | | Fully Connected layer, softmax |

**S2 Table 3.** Representation of DenseNet121 for our classification task of 3D scans - Input size: 256 x 256 x 256

| Layers | Output Size | ResNet50 |
|---|---|---|
| Convolution | 128 x 128 x 128 | 7 x 7 x 7 conv, stride 2 |
| Max Pooling | 64 x 64 x 64 | 3 x 3 x 3 pool, stride 2 |
| Convolutional Layer (type 1) | 64 x 64 x 64 | 1 x 1 x 1 conv -> 3 x 3 x 3 conv -> 1 x 1 x 1 conv -> 1 x 1 x 1 conv |
| Bottleneck | 64 x 64 x 64 | |
| Convolutional Layer (type 2) | 64 x 64 x 64 | 1 x 1 x 1 conv -> 3 x 3 x 3 conv -> 1 x 1 x 1 conv |
| Bottleneck | 64 x 64 x 64 | |

| | | |
|---|---|---|
| Convolutional Layer (type 2) | 64 x 64 x 64 | 1 x 1 x 1 conv -> 3 x 3 x 3 conv -> 1 x 1 x 1 conv |
| Bottleneck | 64 x 64 x 64 | |
| Convolutional Layer (type 1) | 32 x 32 x 32 | 1 x 1 x 1 conv -> 3 x 3 x 3 conv -> 1 x 1 x 1 conv -> 1 x 1 x 1 conv |
| Bottleneck | 32 x 32 x 32 | |
| Convolutional Layer (type 2) | 32 x 32 x 32 | 1 x 1 x 1 conv -> 3 x 3 x 3 conv -> 1 x 1 x 1 conv |
| Bottleneck | 32 x 32 x 32 | |
| Convolutional Layer (type 2) | 32 x 32 x 32 | 1 x 1 x 1 conv -> 3 x 3 x 3 conv -> 1 x 1 x 1 conv |
| Bottleneck | 32 x 32 x 32 | |
| Convolutional Layer (type 2) | 32 x 32 x 32 | 1 x 1 x 1 conv -> 3 x 3 x 3 conv -> 1 x 1 x 1 conv |
| Bottleneck | 32 x 32 x 32 | |
| Convolutional Layer (type 1) | 32 x 32 x 32 | 1 x 1 x 1 conv -> 3 x 3 x 3 conv -> 1 x 1 x 1 conv -> 1 x 1 x 1 conv |
| Bottleneck | 32 x 32 x 32 | |
| Convolutional Layer (type 2) | 32 x 32 x 32 | 1 x 1 x 1 conv -> 3 x 3 x 3 conv -> 1 x 1 x 1 conv |
| Bottleneck | 32 x 32 x 32 | |
| Convolutional Layer (type 2) | 32 x 32 x 32 | 1 x 1 x 1 conv -> 3 x 3 x 3 conv -> 1 x 1 x 1 conv |
| Bottleneck | 32 x 32 x 32 | |
| Convolutional Layer (type 2) | 32 x 32 x 32 | 1 x 1 x 1 conv -> 3 x 3 x 3 conv -> 1 x 1 x 1 conv |
| Bottleneck | 32 x 32 x 32 | |
| Convolutional Layer (type 2) | 32 x 32 x 32 | 1 x 1 x 1 conv -> 3 x 3 x 3 conv -> 1 x 1 x 1 conv |
| Bottleneck | 32 x 32 x 32 | |
| Convolutional Layer (type 1) | 32 x 32 x 32 | 1 x 1 x 1 conv -> 3 x 3 x 3 conv -> 1 x 1 x 1 conv -> 1 x 1 x 1 conv |
| Bottleneck | 32 x 32 x 32 | |
| Convolutional Layer (type 2) | 32 x 32 x 32 | 1 x 1 x 1 conv -> 3 x 3 x 3 conv -> 1 x 1 x 1 conv |
| Bottleneck | 32 x 32 x 32 | |
| Convolutional Layer (type 2) | 32 x 32 x 32 | 1 x 1 x 1 conv -> 3 x 3 x 3 conv -> 1 x 1 x 1 conv |
| Bottleneck | 32 x 32 x 32 | |
| Classification Layer | 1 x 1 x 1 | 7 x 7 x 7 global average pool |

| | |
|---|---|
| | Fully Connected layer, softmax |

**S2 Table 4.** Architecture of ResNet50 - in yellow, the layers for which we extracted the parameters from the pre-trained model Med3d; in light green, the layers for which we continued training the parameters to fine-tune the model and adapt it to the task of predicting Autism.

### S3 - Performance of the models

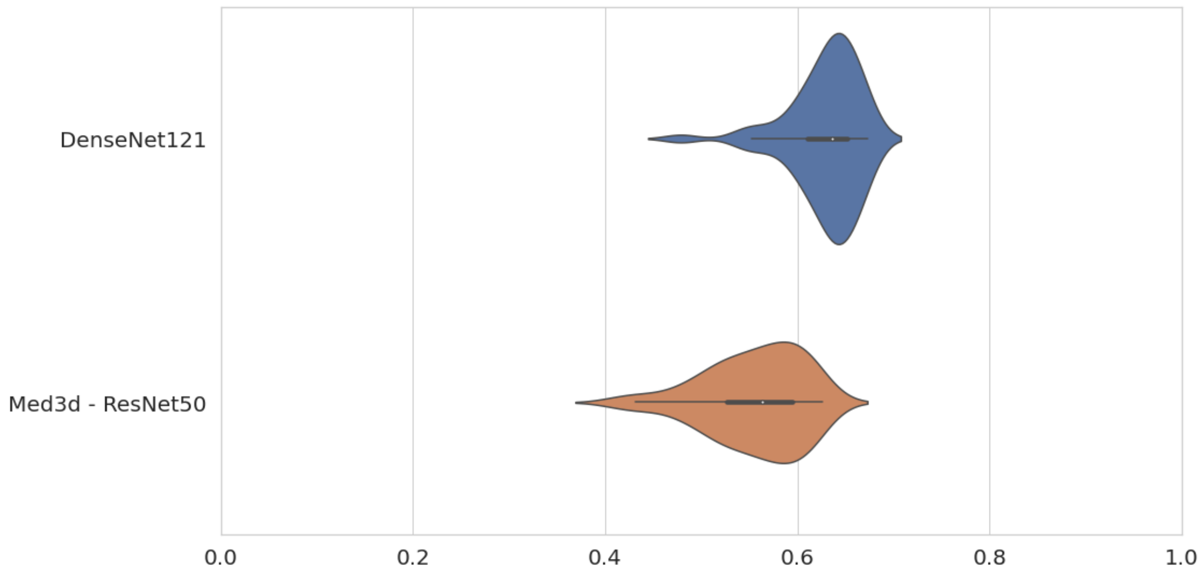


**S3 Fig 1.** Validation set accuracy during training for the two models DenseNet161 and Med3d-ResNet50.

**S3 Fig 1** compares the distributions of validation set accuracies for each model. DenseNet121 tended to have more sSupplemental Table and higher accuracies on the validation set than Med3d-ResNet50.

| | **Med3d - ResNet50 - 42 epochs** | | | **DenseNet121 - 32 epochs** | | | **DenseNet121 - 70 epochs** | | |
|---|---|---|---|---|---|---|---|---|---|
| Subjects | **All** | **Autism** | **non-Autism** | **All** | **Autism** | **on-Autism** | **All** | **Autism** | **on-Autism** |
| **Training set** | Accuracy: 94,2 % ROC AUC: 99,9 % | Accuracy: 85,3 % | Accuracy: 100 % | Accuracy: 65,5 % ROC AUC: 69,1 % | Accuracy: 32,8 % | Accuracy: 86,7% | Accuracy: 69,7 % ROC AUC: 77,1 %: | Accuracy: 68,2 % | Accuracy: 70,8 % |
| **Validation set** | Accuracy: 62,6 % ROC AUC: 62,1 % | Accuracy: 17,6 % | Accuracy: 91,4 % | Accuracy: 66,3 % ROC AUC: 68,8 % | Accuracy: 36,5 % | Accuracy: 85,3 % | Accuracy: 67,4 % ROC AUC: 68,1 % | Accuracy: 66,2 % | Accuracy: 68,1 % |
| **Testing set** | Accuracy: 53,8 % ROC AUC: 57,3 % | Accuracy: 50% | Accuracy: 56,4 % | Accuracy 55,4 %: ROC AUC: 60,7 % | Accuracy: 84,6 % | Accuracy: 35,9 % | Accuracy: 40 % ROC AUC: 38,1 % | Accuracy: 69,2 % | Accuracy: 20,5 % |
| **All the** | Accuracy: 87,7 % | | | Accuracy: 65,2 % | | | Accuracy: 67,9 % | | |

| | | | |
|---|---|---|---|
| **dataset** | ROC AUC: 95,5 % | ROC AUC: 68,4 % | ROC AUC: 74,0 %: |

**S3 Table 5.** Comparison of the performance of the prediction of Autism between the models Med3d - ResNet50 - 42 epochs, DenseNet121 - 32 epochs and DenseNet121 - 70 epochs.

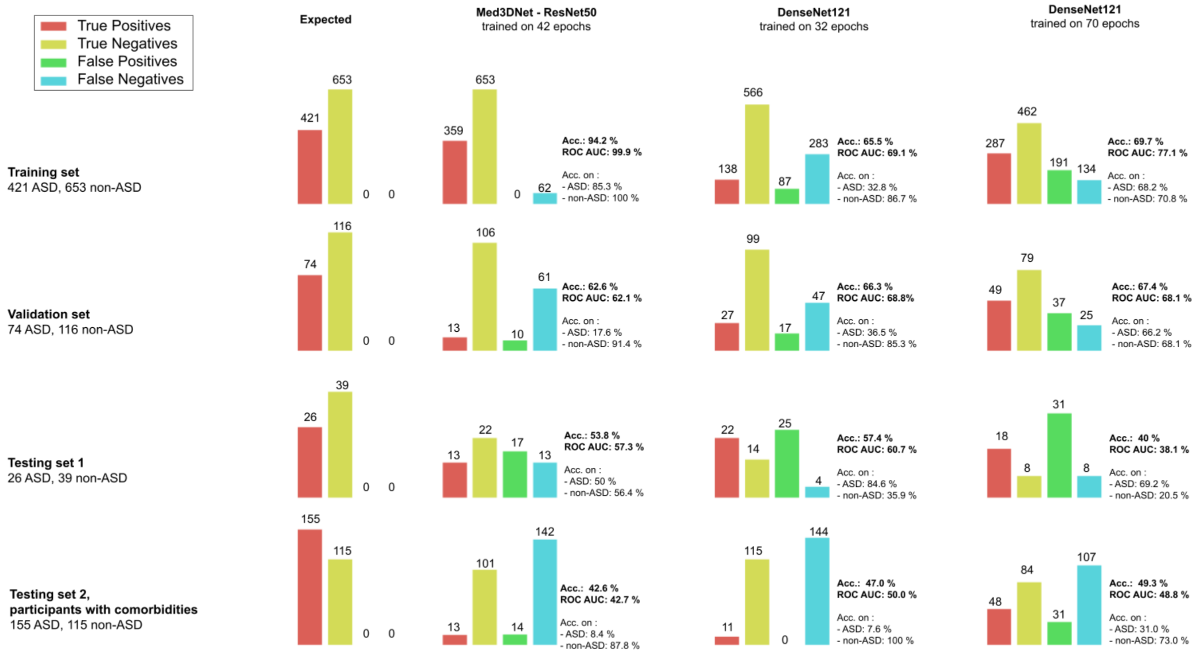


**S3 Fig 2.** True and False Positives and Negatives for each of the three best models - Med3DNet-ResNet50 trained on 42 epochs, DenseNet121 trained on 32 epochs, and DenseNet121 trained on 70 epochs.

**S3 Fig 2** shows the accuracies (in terms of True/False Positives and Negatives) obtained for each of the three best models for prediction of Autism (Autism vs. non-Autism) and each dataset. We can see that Med3d-ResNet50-42ep overfit the data, because the accuracy and ROC AUC scores were very high on the training set (94.2% and 99.9% respectively), but much lower on the validation (acc = 62.6% and AUC = 62.1%) and testing sets (acc = 53.8% and AUC=57.3%). DenseNet121-32ep appeared to be more sSupplemental Table in terms of its overall performance on the training (acc = 65.5% and AUC = 69.1%), validation (acc =66.3% and AUC = 68.8%) and testing (acc =55.4% and AUC = 60.7%) sets.

DenseNet121-70ep had better performance on the training (acc = 69.7% and AUC = 77.1%) and validation (acc = 67.4% and AUC = 68.1%) sets than DenseNet121-32ep, but poorer performance on the testing set (acc = 40% and AUC = 38.1%).

**S3 Fig 2** shows that DenseNet121-32ep has high specificity on the training and validation sets, while having low sensitivity. Paradoxically, it has high sensitivity but low specificity on the testing set. DenseNet121-70ep behaves similarly on the testing set. Nevertheless, on the training and validation sets, we can see that the sensitivity and specificity are balanced and fairly high. Finally, for Med3d-ResNet50-42ep, we observe that the sensitivity and specificity are very high on the training set, are unbalanced on the validation set with a low sensitivity and very high specificity, and are balanced again on the testing set, but with moderate values.

The lowest panel of **S3 Fig 2** shows the accuracies for the second testing set, which included participants with comorbidities. The data show that predicting Autism in the presence of comorbidities is more difficult than predicting Autism when the training and testing sets include only participants without known comorbidities, with a particularly large increase in False Negatives. One potential explanation is that neuroimaging markers become less evident when individuals have another diagnosis involving similar or other neuroimaging markers. Another explanation is that more data are needed to adequately train DL algorithms on the whole spectrum of Autism patients.

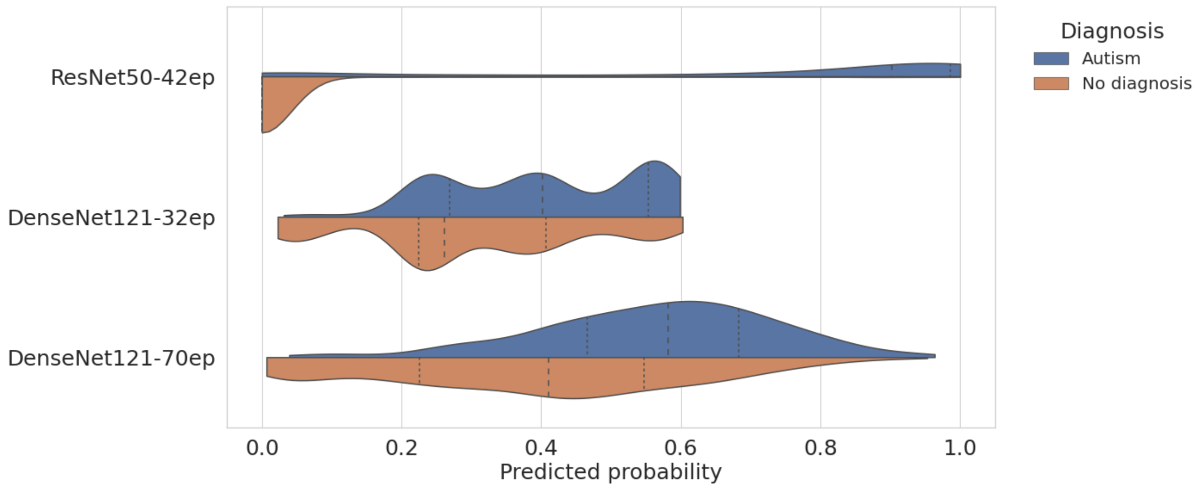


**S3 Fig 3.** Comparison of model predictions across all the datasets without comorbidity (training/validation/test)

**S3 Fig 3** shows that there is a net difference in the distribution of probabilities for Autistic vs non-Autistic participants for the Med3d-ResNet50-42ep model, in line with the observation of overfitting and the very good performance observed for the training set (1074 subjects). For the two other models, the estimated means are distinct, although the distributions overlap. This observation also reflects the accuracy and ROC AUC scores obtained with these two models.

T-tests indicate no significant difference between the age of patients predicted with Autism and the ones predicted with no Autism ($p > .05$).

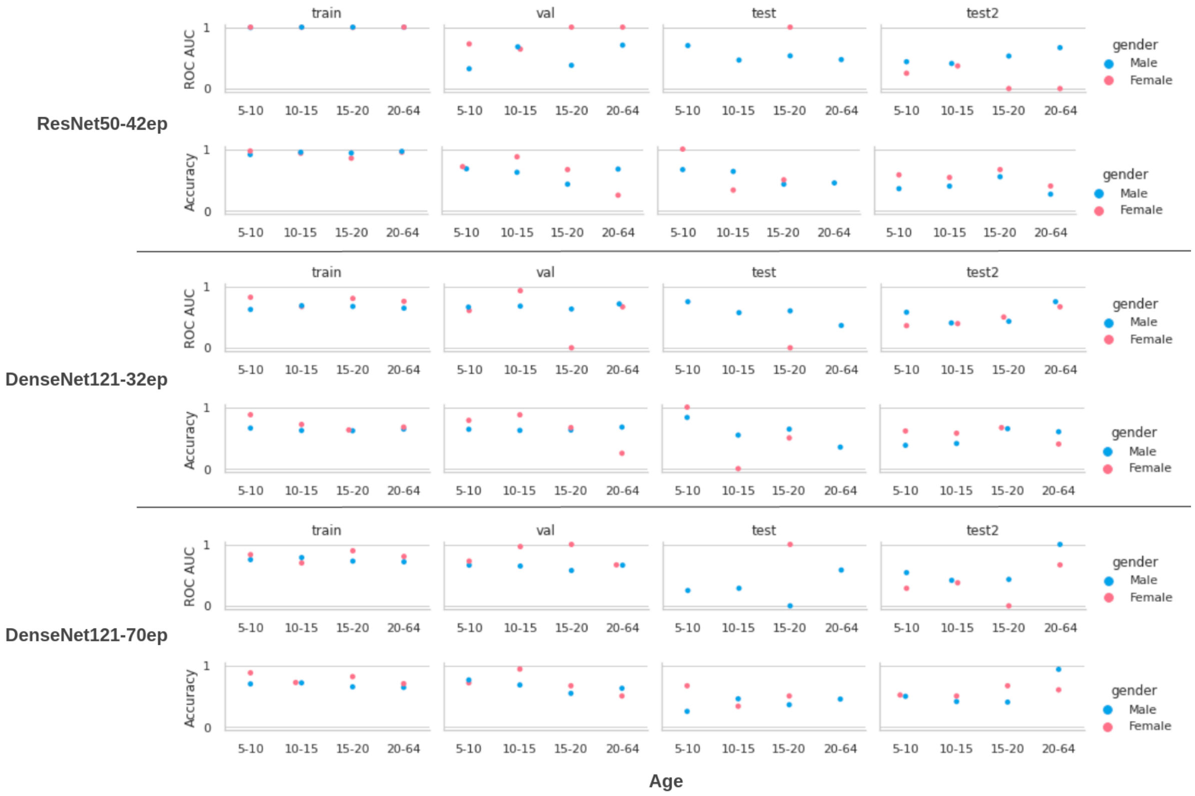


**S3 Fig 4.** ROC AUC and accuracy scores in function of age (between 5 and 10, 10 and 15, 15 and 20, 20 and 64) and gender (male or female) for each model (ResNet50 trained on 42 epochs, DenseNet121 trained on 32 epochs and trained on 70 epochs) for each dataset (training, validation, testing and testing 2 sets).

We observed that the ROC AUC and accuracy scores did not differ between age ranges and between genders in the training set. However, we observed that in the validation set, these scores were variable (see **S3 Fig 4**). We also observed this variation in the two testing sets (see **S3 Fig 4**). This suggests that we should examine the stability of performance between different age ranges and between males and females.

**S4 - Analysis of ADI-R and ADOS scores, age, gender and full IQ**

To better understand differences between the datasets (training, validation and testing sets) and between the classes (Autism and non-Autism), we performed an analysis incorporating the severity scores from ADI-R and ADOS, the age, the gender and the Full IQ scores.

First, we gathered all the information on the diagnosis available in ABIDE I & II. By combining various questionnaires (ADI-R, ADOS Modules 2, 3 and 4), we obtained scores for (1) social interaction (including the Reciprocal Social Interaction Subscore A for ADI-R, the Social Total Subscore of the classic ADOS, the Social Affect Total Subscore for Gotham Algorithm of ADOS, for (2) verbal communication (including the Abnormalities in Communication Subscore (B) for ADI-R, the Communication Total Subscore of the Classic ADOS), (3) for repetitive, restricted or stereotyped behaviors (including the Restricted, Repetitive, and Stereotyped Patterns of Behaviour Subscore (C) for ADI-R, the Stereotyped Behaviours and Restricted Interests Total Subscore of the Classic ADOS, the Restricted and Repetitive Behaviours Total Subscore for Gotham Algorithm of ADOS) and (4) total scores (including the Abnormality of Development Evident at or before 36 months Subscore (D) Total for ADI-R, the Classic ADOS Score, the Gotham Algorithm of ADOS Score) for 452 subjects. Since all of these questionnaires use different scales, we transformed all the scores into Z-scores to compare individuals.

Second, we gathered the predicted class of each patient from each model, and, from it, we created a variable "prediction type" representing the True Positives, False Negatives, True Negatives and False Positives.

Finally, we compared the distributions of Z-scores across prediction types, to investigate whether there were differences in symptom severity scores between True Positives and

False Negatives, and similarly, a difference between True Negatives and False Positives. We visually compared the distribution and performed a T-test

These analyses did not reveal any discernible differences between the predictions of the three models. **S4 Fig 5** illustrates an example of this analysis using Med3d - ResNet50 - 42ep.

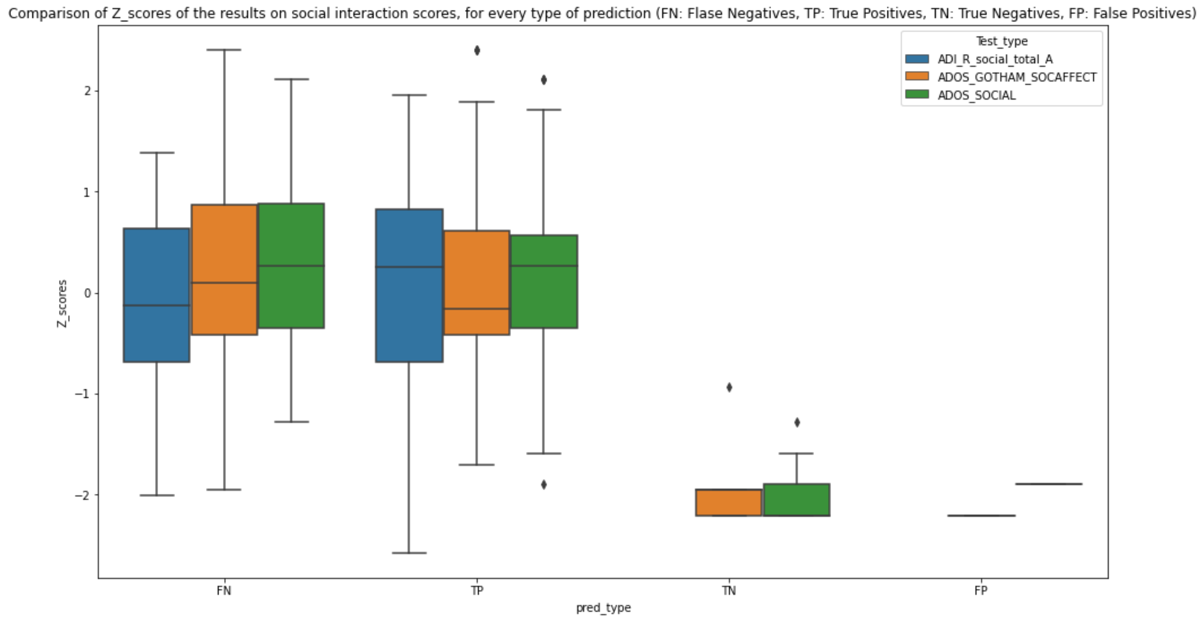


**S4 Fig 5.** Comparison of social interaction Z-scores between False Negatives (FN), True Positives (TP), True Negatives (TN) and False Positives (FP).

We did not find any differences between the three models when we examined the severity scores on the training, validation and testing sets separately. Nor were there differences between males and females in the distribution of probabilities for all the models.

We compared the distribution of age for each prediction type (FN, TP, TN, FP). There was no noticeable difference in age between the samples corresponding respectively to each prediction type for all the models, compared to the distribution of age between the samples corresponding to true labels.

Finally, we also compared the distribution of Full IQ score for each prediction type (FN, TP, TN, FP). There was no noticeable difference between the samples corresponding respectively to each prediction type or all the models, compared to the distribution of FIQ between the samples corresponding to true labels.

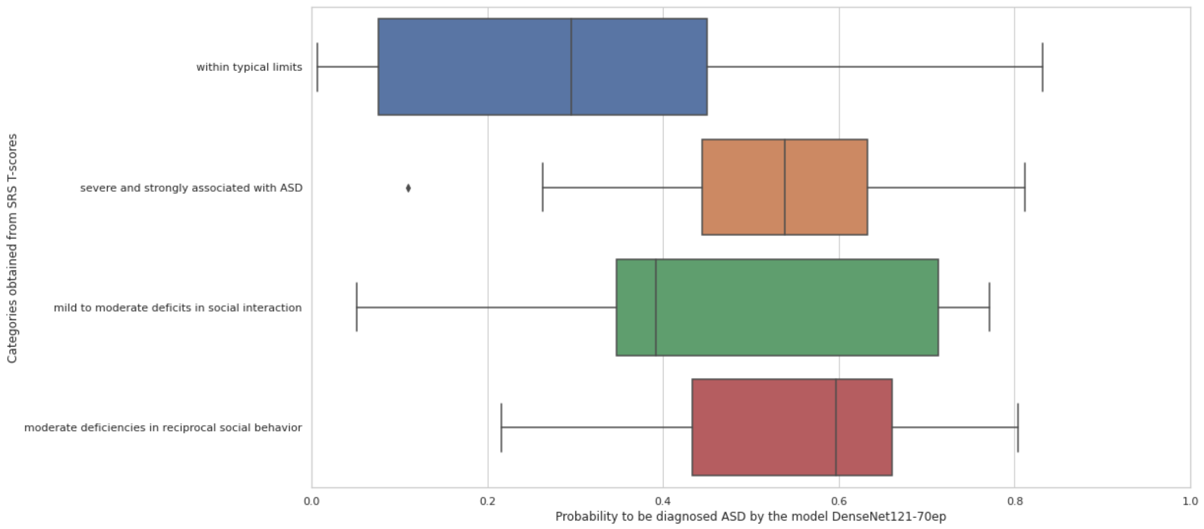


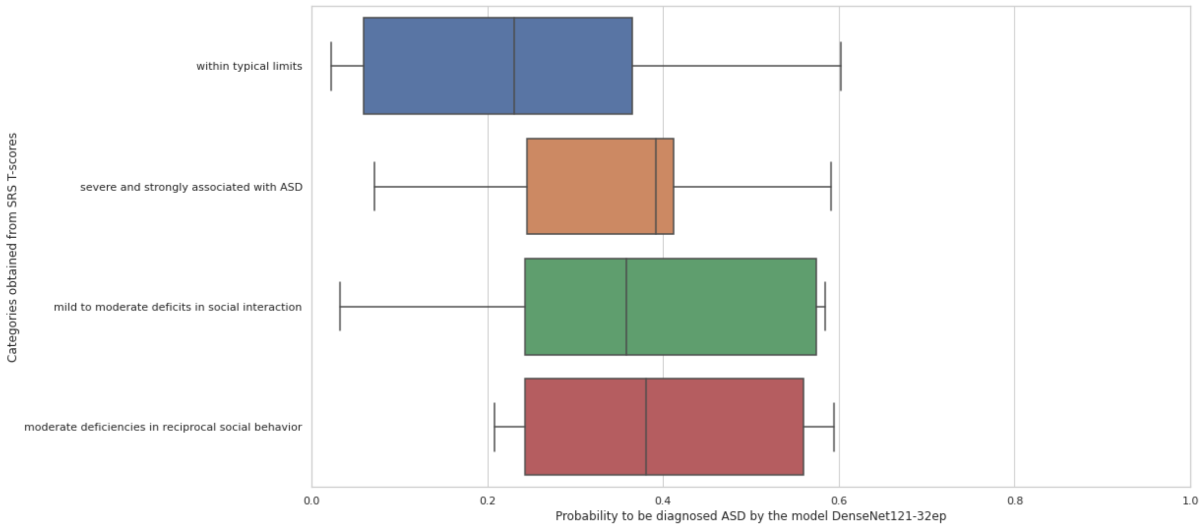

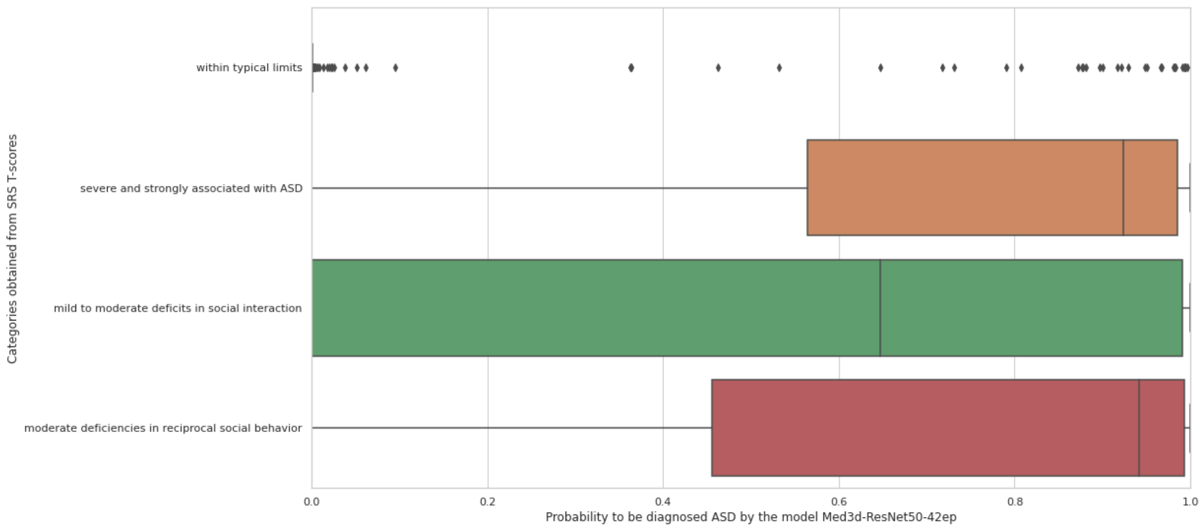


**S4 Fig 6.** Probability scores of each model per category obtained from SRS T-scores in ABIDE 2

In **S4 Fig 6**, for every model, the distribution of probability scores is shown for categories created on the basis of the total T-scores of the SRS-2. “Within typical limits” corresponds to a T-score lower than 59, “mild to moderate difficulties in social interaction” corresponds to a T-score between 60 and 65, “moderate difficulties in reciprocal social behaviour” corresponds to a T-score between 66 and 75, and “severe difficulties, strongly associated with Autism” to a T-score greater than 76. We observed that DenseNet121-70ep had a distribution of probability scores that was consistent with these severity scores, with the majority of scores lower than 0.5 for the category “within typical limits”, and the majority of scores greater than 0.5 or the three other categories.

### S5 - Most important regions for the prediction of True Positives

| | | | R42 | | D32 | | D70 | |
|---|---|---|---|---|---|---|---|---|
| | | | No comorb | With comorb | No comorb | With comorb | No comorb | With comorb |
| Frontal lobe | Left | Frontal operculum | 2 | 2 | 0 | 0 | 2 | 2 |
| Frontal lobe | Left | Middle frontal gyrus | 3 | 4 | 0 | 0 | 0 | 0 |
| Frontal lobe | Left | Precentral gyrus medial segment | 0 | 0 | 3 | 4 | 1 | 2 |
| Frontal lobe | Left | Precentral gyrus | 1 | 1 | 0 | 0 | 2 | 3 |
| Frontal lobe | Left | Triangular part of the inferior frontal gyrus | 3 | 4 | 0 | 0 | 1 | 1 |
| Limbic system and associated structures | Left | Anterior Cingulate Gyrus | 3 | 4 | 0 | 0 | 0 | 0 |
| Limbic system and associated structures | Left | Parahippocampal gyrus | 1 | 2 | 0 | 0 | 3 | 4 |
| Limbic system and associated structures | Left | Subcallosal area | 3 | 4 | 0 | 0 | 0 | 1 |
| Parietal lobe | Left | Central operculum | 2 | 2 | 0 | 0 | 1 | 2 |
| Parietal lobe | Left | Parietal operculum | 0 | 1 | 2 | 3 | 2 | 3 |
| Parietal lobe | Left | Parietal white matter | 0 | 0 | 2 | 2 | 2 | 3 |
| Parietal lobe | Left | Supplementary motor cortex | 0 | 0 | 1 | 2 | 1 | 2 |
| Parietal lobe | Left | Supramarginal gyrus | 0 | 0 | 2 | 3 | 2 | 3 |
| occipital lobe | Left | Posterior orbital gyrus | 2 | 3 | 0 | 0 | 2 | 2 |
| subcortical structures | Right | Ventral diencephalon | 0 | 1 | 2 | 3 | 1 | 1 |
| temporal lobe | Left | Planum temporale | 1 | 1 | 1 | 2 | 2 | 3 |
| temporal lobe | Left | Superior temporal gyrus | 2 | 2 | 0 | 0 | 2 | 2 |
| temporal lobe | Left | Temporal pole | 2 | 2 | 0 | 0 | 3 | 3 |

| | | | | | | | | |
|---|---|---|---|---|---|---|---|---|
| **temporal lobe** | **Left** | **Transverse temporal gyrus** | **1** | **1** | **0** | **1** | **1** | **2** |

**S5 Table 6.** Best regions for predicting True Positives (TP, i.e. true diagnosis of Autism): each row is for one region, each column is for one model (R42 for ResNet50 trained on 42 epochs, D32 for DenseNet121 trained on 32 epochs, D70 for DenseNet121 trained on 70 epochs) and one combination of datasets considered (training+validation+testing 1 sets ("no comorb" for no comorbidity), or all these sets + testing set 2 ("with comorb" for containing subjects with comorbidities), each case returns the number of datasets where the region was important for predicting TP for the model considered.

**S5 Table 6** summarizes the most important regions for predicting TP, which identified regions that were common across models. Limiting the results in this way enables us to reduce the bias effect of each algorithm (that leads to regions important only for one model). We used a methodology analogous to a traditional Machine Learning pipeline here, which identified features on the basis of their importance.

**S5 Table 6** gives us different types of information:

- The most replicable regions for predicting TP between the models
- The replicability of the regions found between the datasets not containing subjects with comorbidity (thanks to the number in each case in the columns "Training + val. + testing 1 sets")
- The replicability of the regions found between datasets without comorbidity and dataset with comorbidity (thanks to the number in each case in the columns "Training + val. + testing 1 & 2 sets"). This is also shown by the changes highlighted in light red.

For instance, for the model Med3D-ResNet50 trained on 42 epochs, we found that Right-ACgG-anterior-cingulate-gyrus is an important region for predicting TP on three over the three datasets into the datasets without comorbidity, and on four over the four datasets

“Training + val. + testing 1 & 2 sets” that contains subjects with comorbidities in testing set 2. Thus, for the model Med3D-ResNet50 trained on 42 epochs, Right-ACgG-anterior-cingulate-gyrus is important for the prediction of TP, and, by extension, for the detection of Autism, and was robust to comorbidities.

In **S5 Table 6**, we observed that several regions were important for the three models, including Left Planum Temporale, Left Parietal Operculum, Right Ventral Diencephalon. However, we saw that for the four regions the replicability is low between the datasets.

We also noticed that, on the one hand, a lot of regions were commonly important for the two DenseNet models but not for ResNet50, including Left Supramarginal Gyrus, Left Parietal White Matter and Left precentral gyrus medial segment. On the other hand, Left subcallosal area, Left Middle Frontal gyrus, Left-MFC-medial-frontal-cortex and Left anterior cingulate gyrus were important for ResNet50 but not for the two DenseNet models, and their importance replicated well over the datasets, including the one with comorbidities.

Further, several regions were important to both ResNet50 and to DenseNet121-70ep, including Left triangular part of the inferior frontal gyrus, Left Temporal Pole , Left Precentral Gyrus, Left posterior orbital gyrus, Left Parahippocampal gyrus. We noticed that for Left triangular part of the inferior frontal gyrus, the replicability over the datasets without comorbidity was higher for the ResNet50 model than for the DenseNet121-70ep model, while we observed the opposite for Left Parahippocampal gyrus. However, we noticed that for the ResNet50 model, the importance of Left Precentral Gyrus did not replicate in the testing set 2 with comorbidities whereas for DenseNet121-70ep it did. The converse was observed for Left posterior orbital gyrus. This disparity is illustrative of the bias introduced by each model, due to the different architectures and levels of optimisation. Even though we set the optimizer parameters similarly between the models, due to the inherent difference in the designs, the models tend to approximate a function that achieves the task of detecting

Autism in different ways. This also underlines the importance of considering different types of models in Deep Learning when possible (machine/funding limitation), analogously to more traditional Machine Learning pipelines of analysis.

With regard to participants with comorbidities, we see from **S5 Table 6** that all the regions important for ResNet50-42ep, but which were not shared with the other models, replicated well in the test set with comorbidities. Globally, the models ResNet50-42ep and DenseNet70-70ep have an equivalent number of areas that were important for predicting TP and which replicated well in the testing set 2, higher than for the model DenseNet121-32ep. Another interesting point is that certain regions that were not among the most important for predicting TP in the datasets without comorbidities appear important for predicting TP in the dataset with subjects who did have comorbidities. This includes Left subcallosal area for DenseNet121-70ep, and Right Ventral Diencephalon, Left Parietal Operculum for ResNet50-42ep.

Summarizing the Supplemental Table, and taking each model separately, the most important regions for predicting Autism across all the models and between datasets are Left triangular part of the inferior frontal gyrus, Left subcallosal area, Left Parahippocampal gyrus, Left precentral gyrus medial segment, Left Middle Frontal gyrus and Left anterior cingulate gyrus.

On the one hand, this result can help us identify neuroimaging markers of Autism, by combining the findings between the models, using Deep Learning as a way to extract feature importance in a manner similar to Random Forest, for instance. On the other hand, this shows that each model tends to focus on specific parts in the brain, capturing different patterns than the other models, making it difficult to select one model that works best.

**S6 - Most important regions for the prediction of True Negatives**

Overall, after aggregating all the datasets, among the 79 areas most important for predicting TN, 10 areas combining left and right hemispheres, 24 in the left hemisphere and 2 in the right hemisphere were commonly predictive for TP.

Keeping only the areas that replicated the most over the datasets, the areas predictive for TN were largely different from the ones that were important for TP.

| | | | R42 | | D32 | | D70 | |
|---|---|---|---|---|---|---|---|---|
| | | | No comorb | With comorb | No comorb | With comorb | No comorb | With comorb |
| Frontal lobe | Left | Frontal operculum | 3 | 4 | 0 | 0 | 0 | 0 |
| Limbic system and associated structures | Left | Posterior cingulate gyrus | 1 | 1 | 2 | 3 | 1 | 1 |
| Limbic system and associated structures | Right | Posterior cingulate gyrus | 0 | 0 | 2 | 3 | 1 | 1 |
| Parietal lobe | Left | Precuneus | 0 | 0 | 2 | 3 | 1 | 2 |
| Parietal lobe | Left | Superior parietal lobule | 0 | 0 | 2 | 3 | 2 | 3 |
| cerebellum | None | Vermal Lobules VI-VII | 0 | 0 | 3 | 4 | 2 | 2 |
| cerebellum | Left | Cerebellum exterior | 0 | 0 | 2 | 3 | 2 | 2 |
| occipital lobe | Left | Angular gyrus | 0 | 0 | 2 | 3 | 2 | 3 |
| occipital lobe | Left | Calcarine cortex | 0 | 0 | 0 | 1 | 2 | 3 |
| occipital lobe | Left | Cuneus | 0 | 0 | 1 | 1 | 2 | 3 |
| occipital lobe | Left | Inferior occipital | 0 | 0 | 3 | 4 | 2 | 3 |

| | | | | | | | | |
|---|---|---|---|---|---|---|---|---|
| | | gyrus | | | | | | |
| occipital lobe | Left | Lingual gyrus | 0 | 0 | 2 | 3 | 2 | 3 |
| occipital lobe | Left | Middle occipital gyrus | 0 | 0 | 2 | 2 | 2 | 3 |
| occipital lobe | Left | Occipital fusiform gyrus | 0 | 0 | 3 | 4 | 2 | 3 |
| occipital lobe | Left | Occipital White Matter | 0 | 0 | 2 | 3 | 2 | 3 |
| occipital lobe | Left | Superior occipital gyrus | 0 | 0 | 1 | 2 | 3 | 4 |
| subcortical structures | Left | Thalamus | 3 | 4 | 0 | 0 | 0 | 0 |
| subcortical structures | Right | Ventral diencephalon | 2 | 3 | 0 | 0 | 1 | 1 |
| temporal lobe | Left | Middle temporal gyrus | 0 | 0 | 2 | 2 | 2 | 2 |
| temporal lobe | Left | Planum polare | 3 | 4 | 0 | 0 | 0 | 0 |

**S6 Table 7.** Best regions for predicting True Negatives (TN, i.e. no diagnosis of Autism): each row is for one region, each column is for one model and one combination of datasets considered (training+validation+testing 1 sets (no comorbidity), or all these sets + testing set 2 (containing subjects with comorbidities)), each case returns the number of datasets where the region was important for predicting TN for the model considered.

### S7- Most replicable regions for *False Positives* and *False Negatives*

| | | | R42 | | D32 | | D70 | |
|---|---|---|---|---|---|---|---|---|
| | | | No comorb | With comorb | No comorb | With comorb | No comorb | With comorb |
| Frontal lobe | Left | Frontal operculum | 1 | 1 | 0 | 0 | 3 | 3 |
| Frontal lobe | Left | Middle frontal gyrus | 1 | 2 | 0 | 0 | 2 | 3 |
| Frontal lobe | Left | Precentral gyrus medial segment | 0 | 0 | 2 | 2 | 2 | 3 |
| Frontal lobe | Left | Precentral gyrus | 1 | 2 | 0 | 0 | 3 | 3 |
| Frontal lobe | Left | Triangular part of the inferior frontal gyrus | 1 | 1 | 0 | 0 | 3 | 3 |
| Frontal lobe | Right | Precentral gyrus medial segment | 0 | 0 | 3 | 3 | 0 | 1 |
| Parietal lobe | Left | Parietal operculum | 1 | 1 | 2 | 2 | 1 | 1 |
| Parietal lobe | Left | Parietal white matter | 0 | 0 | 2 | 2 | 2 | 2 |
| Parietal lobe | Left | Supplementary motor cortex | 0 | 0 | 3 | 3 | 1 | 2 |
| Parietal lobe | Left | Supramarginal gyrus | 0 | 1 | 2 | 2 | 2 | 3 |
| Parietal lobe | Left | Superior parietal lobule | 0 | 0 | 3 | 3 | 1 | 2 |
| occipital lobe | Left | Angular gyrus | 0 | 0 | 3 | 3 | 2 | 2 |
| occipital lobe | Left | Posterior orbital gyrus | 2 | 2 | 0 | 0 | 3 | 3 |
| temporal lobe | Left | Postcentral gyrus | 0 | 0 | 2 | 2 | 2 | 2 |
| temporal lobe | Left | Temporal pole | 1 | 1 | 0 | 0 | 3 | 3 |

**S7 Table 8.** Best regions for predicting False Positives (FP, i.e. prediction of Autism whereas no diagnosis Autism): each row is for one region, each column is for one model and one combination of datasets considered (training+validation+testing 1 sets (no comorbidity), or all these sets + testing set 2 (containing subjects with comorbidities)), each case returns the number of datasets where the region was important for predicting TN for the model considered.

| | | | R42 | | D32 | | D70 | |
|---|---|---|---|---|---|---|---|---|
| | | | No comorb | With comorb | No comorb | With comorb | No comorb | With comorb |
| **Frontal lobe** | **Left** | **Frontal operculum** | 3 | 4 | 0 | 0 | 0 | 0 |
| **Limbic system and associated structures** | **Left** | **Posterior cingulate gyrus** | 0 | 1 | 2 | 3 | 1 | 2 |
| **Limbic system and associated structures** | **Right** | **Posterior cingulate gyrus** | 0 | 1 | 2 | 3 | 1 | 1 |
| **Parietal lobe** | **Left** | **Precuneus** | 0 | 0 | 3 | 4 | 1 | 2 |
| **Parietal lobe** | **Left** | **Superior parietal lobule** | 0 | 0 | 1 | 2 | 1 | 2 |
| **cerebellum** | **None** | **Vermal Lobules VII-X** | 0 | 0 | 1 | 2 | 1 | 2 |
| **cerebellum** | **Left** | **Cerebellum exterior** | 0 | 0 | 2 | 3 | 1 | 2 |
| **occipital lobe** | **Left** | **Angular gyrus** | 0 | 0 | 2 | 3 | 2 | 3 |
| **occipital lobe** | **Left** | **Cuneus** | 0 | 0 | 1 | 1 | 2 | 3 |
| **occipital lobe** | **Left** | **Inferior occipital gyrus** | 0 | 0 | 3 | 4 | 2 | 3 |
| **occipital lobe** | **Left** | **Lingual gyrus** | 0 | 0 | 2 | 3 | 2 | 3 |
| **occipital lobe** | **Left** | **Middle occipital gyrus** | 0 | 0 | 2 | 3 | 2 | 3 |
| **occipital lobe** | **Left** | **Occipital fusiform gyrus** | 0 | 0 | 3 | 4 | 1 | 2 |

| | | | | | | | | |
|---|---|---|---|---|---|---|---|---|
| **occipital lobe** | **Left** | **Occipital White Matter** | **0** | **0** | **2** | **3** | **2** | **3** |
| **occipital lobe** | **Left** | **Superior occipital gyrus** | **0** | **0** | **1** | **1** | **2** | **3** |
| **temporal lobe** | **Left** | **Middle temporal gyrus** | **0** | **1** | **2** | **2** | **2** | **2** |
| **temporal lobe** | **Left** | **Planum polare** | **3** | **4** | **0** | **0** | **0** | **0** |
| **temporal lobe** | **Left** | **Superior temporal gyrus** | **3** | **4** | **0** | **0** | **0** | **0** |

**S7 Table 9.** Best regions for predicting False Negatives (FN, i.e. no prediction of Autism whereas diagnosed Autism): each row is for one region, each column is for one model and one combination of datasets considered (training+validation+testing 1 sets (no comorbidity), or all these sets + testing set 2 (containing subjects with comorbidities)), each case returns the number of datasets where the region was important for predicting TN for the model considered.

### S8 - True Positives by Gender

| | | | R42 | | D32 | | D70 | |
|---|---|---|---|---|---|---|---|---|
| | | | No comorb | With comorb | No comorb | With comorb | No comorb | With comorb |
| Frontal lobe | Left | Frontal operculum | 2 | 2 | 0 | 0 | 2 | 3 |
| Frontal lobe | Left | Middle frontal gyrus | 3 | 4 | 0 | 0 | 0 | 0 |
| Frontal lobe | Left | Precentral gyrus medial segment | 0 | 0 | 3 | 4 | 1 | 2 |
| Frontal lobe | Left | Precentral gyrus | 1 | 1 | 0 | 0 | 3 | 4 |
| Frontal lobe | Left | Triangular part of the inferior frontal gyrus | 3 | 4 | 0 | 0 | 1 | 1 |
| Limbic system and associated structures | Left | Anterior Cingulate Gyrus | 3 | 4 | 0 | 0 | 0 | 0 |
| Limbic system and associated structures | Left | Parahippocampal gyrus | 1 | 2 | 1 | 1 | 3 | 4 |
| Limbic system and associated structures | Left | Posterior insula | 2 | 2 | 0 | 0 | 2 | 2 |
| Limbic system and associated structures | Left | Subcallosal area | 2 | 3 | 0 | 0 | 1 | 2 |
| Parietal lobe | Left | Central operculum | 2 | 2 | 0 | 0 | 2 | 3 |
| Parietal lobe | Left | Parietal operculum | 1 | 2 | 1 | 2 | 2 | 3 |
| Parietal lobe | Left | Supramarginal gyrus | 0 | 0 | 2 | 3 | 2 | 3 |
| occipital lobe | Left | Posterior orbital gyrus | 2 | 3 | 0 | 0 | 2 | 2 |
| subcortical structures | Right | Ventral diencephalon | 0 | 1 | 2 | 3 | 1 | 1 |
| temporal lobe | Left | Planum temporale | 1 | 1 | 1 | 2 | 2 | 3 |
| temporal lobe | Left | Superior temporal gyrus | 2 | 2 | 0 | 0 | 2 | 2 |
| temporal lobe | Left | Temporal pole | 2 | 2 | 0 | 0 | 3 | 3 |
| temporal lobe | Left | Transverse temporal gyrus | 1 | 1 | 1 | 2 | 1 | 2 |

**S8 Table 10.** Best regions for predicting True Positives (TP, i.e. true diagnosis of Autism) for Boys

| | | | R42 | | D32 | | D70 | |
|---|---|---|---|---|---|---|---|---|
| | | | No comorb | With comorb | No comorb | With comorb | No comorb | With comorb |
| Frontal lobe | Left | Triangular part of the inferior frontal gyrus | 1 | 2 | 1 | 1 | 1 | 1 |
| Frontal lobe | Right | Precentral gyrus medial segment | 0 | 0 | 2 | 3 | 0 | 1 |
| Limbic system and associated structures | Left | Posterior cingulate gyrus | 0 | 0 | 2 | 3 | 2 | 3 |
| Limbic system and associated structures | Right | Middle cingulate gyrus | 0 | 1 | 0 | 1 | 1 | 2 |
| Parietal lobe | Left | Parietal operculum | 1 | 1 | 1 | 2 | 2 | 3 |
| Parietal lobe | Left | Parietal white matter | 0 | 0 | 0 | 1 | 2 | 3 |
| Parietal lobe | Left | Supramarginal gyrus | 0 | 0 | 1 | 2 | 2 | 3 |
| Parietal lobe | Left | Superior parietal lobule | 0 | 0 | 2 | 2 | 3 | 3 |
| Parietal lobe | Right | Supplementary motor cortex | 1 | 1 | 0 | 1 | 1 | 2 |
| occipital lobe | Left | Angular gyrus | 0 | 0 | 2 | 2 | 2 | 2 |
| occipital lobe | Left | Occipital pole | 0 | 0 | 1 | 2 | 1 | 2 |
| temporal lobe | Left | Postcentral gyrus medial segment | 0 | 1 | 2 | 2 | 2 | 2 |
| temporal lobe | Left | Postcentral gyrus | 0 | 0 | 0 | 1 | 2 | 3 |
| temporal lobe | Right | Postcentral gyrus medial segment | 0 | 0 | 2 | 2 | 2 | 2 |

**S8 Table 11.** Best regions for predicting True Positives (TP, i.e. true diagnosis of Autism) for Girls.

### S9 - True Positives by Gender and Age

| | | | R42 | | D32 | | D70 | |
|---|---|---|---|---|---|---|---|---|
| | | | No comorb | With comorb | No comorb | With comorb | No comorb | With comorb |
| Frontal lobe | Left | Middle frontal gyrus | 2 | 2 | 0 | 0 | 2 | 2 |
| Frontal lobe | Left | Precentral gyrus | 2 | 2 | 0 | 0 | 3 | 4 |
| Frontal lobe | Left | Triangular part of the inferior frontal gyrus | 1 | 2 | 0 | 0 | 2 | 3 |
| Limbic system and associated structures | Left | Hippocampus | 2 | 2 | 0 | 0 | 1 | 2 |
| Parietal lobe | Left | Central operculum | 2 | 2 | 0 | 0 | 3 | 4 |
| Parietal lobe | Left | Supramarginal gyrus | 1 | 2 | 0 | 0 | 1 | 2 |
| occipital lobe | Left | Posterior orbital gyrus | 1 | 1 | 0 | 0 | 3 | 4 |
| temporal lobe | Left | Temporal pole | 2 | 2 | 0 | 0 | 2 | 3 |

**S9 Table 12.** Best regions for predicting True Positives (TP, i.e. true diagnosis of Autism) for Boys aged 5 to 10.

| | | | R42 | | D32 | | D70 | |
|---|---|---|---|---|---|---|---|---|
| | | | No comorb | With comorb | No comorb | With comorb | No comorb | With comorb |
| Frontal lobe | Left | Frontal White Matter | 1 | 1 | 0 | 0 | 2 | 3 |
| Frontal lobe | Left | Triangular part of the inferior frontal gyrus | 3 | 4 | 0 | 0 | 2 | 2 |
| Frontal lobe | Right | Precentral gyrus medial segment | 0 | 0 | 3 | 3 | 1 | 1 |
| Limbic system and associated structures | Left | Parahippocampal gyrus | 0 | 1 | 1 | 2 | 2 | 3 |

| | | | | | | | | |
|---|---|---|---|---|---|---|---|---|
| **Limbic system and associated structures** | **Left** | **Subcallosal area** | **3** | **4** | **0** | **0** | **0** | **0** |
| **Parietal lobe** | **Left** | **Parietal operculum** | **1** | **2** | **0** | **1** | **1** | **2** |
| **Parietal lobe** | **Left** | **Supramarginal gyrus** | **0** | **0** | **3** | **4** | **1** | **2** |
| **Parietal lobe** | **Left** | **Superior parietal lobule** | **0** | **0** | **3** | **3** | **1** | **1** |
| **occipital lobe** | **Left** | **Posterior orbital gyrus** | **2** | **3** | **0** | **0** | **3** | **3** |
| **temporal lobe** | **Left** | **Planum temporale** | **1** | **1** | **0** | **1** | **1** | **2** |
| **temporal lobe** | **Left** | **Postcentral gyrus** | **0** | **0** | **1** | **2** | **1** | **2** |
| **temporal lobe** | **Left** | **Temporal pole** | **2** | **3** | **0** | **0** | **3** | **3** |
| **temporal lobe** | **Left** | **Transverse temporal gyrus** | **2** | **2** | **1** | **2** | **2** | **3** |

**S9 Table 13.** Best regions for predicting True Positives (TP, i.e. true diagnosis of Autism) for Boys aged 10 to 15.

| | | | **R42** | | **D32** | | **D70** | |
|---|---|---|---|---|---|---|---|---|
| | | | **No comorb** | **With comorb** | **No comorb** | **With comorb** | **No comorb** | **With comorb** |
| **Frontal lobe** | **Left** | **Precentral gyrus medial segment** | **0** | **0** | **2** | **3** | **2** | **3** |
| **Limbic system and associated structures** | **Left** | **Posterior cingulate gyrus** | **0** | **0** | **2** | **2** | **2** | **2** |
| **Limbic system and associated structures** | **Left** | **Parahippocampa l gyrus** | **1** | **2** | **1** | **1** | **1** | **1** |
| **Limbic system and associated structures** | **Right** | **Cingulate White Matter** | **1** | **2** | **0** | **1** | **2** | **2** |
| **Limbic system and associated structures** | **Right** | **Middle cingulate gyrus** | **0** | **0** | **1** | **2** | **1** | **2** |
| **Parietal lobe** | **Left** | **Parietal operculum** | **1** | **2** | **2** | **3** | **1** | **1** |
| **Parietal lobe** | **Left** | **Parietal white** | **0** | **0** | **2** | **2** | **2** | **2** |

| | | | | | | | | |
|---|---|---|---|---|---|---|---|---|
| | | matter | | | | | | |
| Parietal lobe | Left | Supplementary motor cortex | 0 | 0 | 2 | 3 | 1 | 2 |
| Parietal lobe | Left | Supramarginal gyrus | 0 | 0 | 2 | 3 | 2 | 2 |
| Parietal lobe | Right | Supplementary motor cortex | 0 | 0 | 1 | 2 | 2 | 3 |
| occipital lobe | Left | Angular gyrus | 0 | 0 | 2 | 2 | 2 | 2 |
| occipital lobe | Left | Posterior orbital gyrus | 3 | 3 | 0 | 0 | 1 | 1 |
| subcortical structures | Left | Putamen | 3 | 3 | 0 | 0 | 1 | 1 |
| subcortical structures | Right | Ventral diencephalon | 0 | 1 | 2 | 3 | 1 | 2 |
| temporal lobe | Left | Postcentral gyrus | 0 | 0 | 0 | 1 | 2 | 3 |
| temporal lobe | Left | Superior temporal gyrus | 2 | 3 | 0 | 0 | 1 | 1 |
| temporal lobe | Left | Temporal pole | 2 | 2 | 0 | 0 | 1 | 2 |

**S9 Table 14.** Best regions for predicting True Positives (TP, i.e. true diagnosis of Autism) for Boys aged 15 to 20.

| | | | R42 | | D32 | | D70 | |
|---|---|---|---|---|---|---|---|---|
| | | | No comorb | With comorb | No comorb | With comorb | No comorb | With comorb |
| Frontal lobe | Left | Precentral gyrus medial segment | 0 | 1 | 2 | 2 | 0 | 1 |
| Frontal lobe | Left | Opercular part of the inferior frontal gyrus | 2 | 3 | 0 | 0 | 1 | 2 |
| Frontal lobe | Left | Precentral gyrus | 2 | 2 | 0 | 0 | 1 | 2 |
| Limbic system and associated structures | Left | Parahippocampal gyrus | 0 | 0 | 1 | 1 | 3 | 3 |
| Limbic system and associated structures | Left | Posterior insula | 2 | 2 | 0 | 0 | 2 | 2 |

| | | | | | | | | |
|---|---|---|---|---|---|---|---|---|
| **Limbic system and associated structures** | **Left** | **Subcallosal area** | **2** | **2** | **0** | **0** | **1** | **2** |
| **Parenchyma** | **None** | **3rd Ventricle** | **0** | **1** | **1** | **1** | **1** | **2** |
| **Parietal lobe** | **Left** | **Parietal operculum** | **0** | **1** | **2** | **2** | **2** | **2** |
| **Parietal lobe** | **Left** | **Supplementary motor cortex** | **0** | **1** | **1** | **1** | **1** | **2** |
| **subcortical structures** | **Right** | **Ventral diencephalon** | **0** | **0** | **3** | **3** | **1** | **1** |
| **temporal lobe** | **Left** | **Planum temporale** | **1** | **2** | **1** | **1** | **2** | **2** |
| **temporal lobe** | **Left** | **Superior temporal gyrus** | **2** | **2** | **0** | **0** | **3** | **3** |
| **temporal lobe** | **Left** | **Temporal pole** | **2** | **2** | **0** | **0** | **3** | **3** |

**S9 Table 15.** Best regions for predicting True Positives (TP, i.e. true diagnosis of Autism) for Boys aged 20 to 64.

| | | | **R42** | | **D32** | | **D70** | |
|---|---|---|---|---|---|---|---|---|
| | | | **No comorb** | **With comorb** | **No comorb** | **With comorb** | **No comorb** | **With comorb** |
| **Limbic system and associated structures** | **Left** | **Parahippocam pal gyrus** | **1** | **1** | **1** | **1** | **1** | **1** |
| **Parietal lobe** | **Left** | **Parietal white matter** | **0** | **0** | **1** | **1** | **2** | **2** |
| **Parietal lobe** | **Left** | **Supramargina l gyrus** | **0** | **0** | **1** | **1** | **2** | **2** |
| **Parietal lobe** | **Left** | **Superior parietal lobule** | **0** | **0** | **1** | **1** | **2** | **2** |
| **occipital lobe** | **Left** | **Angular gyrus** | **0** | **0** | **1** | **1** | **2** | **2** |
| **subcortical structures** | **Left** | **Thalamus** | **1** | **1** | **1** | **1** | **1** | **1** |

**S9 Table 16.** Best regions for predicting True Positives (TP, i.e. true diagnosis of Autism) for Girls aged 5 to 10.

| | | | R42 | | D32 | | D70 | |
|---|---|---|---|---|---|---|---|---|
| | | | No comorb | With comorb | No comorb | With comorb | No comorb | With comorb |
| **Frontal lobe** | **Left** | **Precentral gyrus medial segment** | 0 | 0 | 1 | 2 | 1 | 2 |
| **Frontal lobe** | **Right** | **Precentral gyrus medial segment** | 0 | 0 | 1 | 2 | 1 | 2 |
| **Limbic system and associated structures** | **Left** | **Posterior cingulate gyrus** | 0 | 0 | 1 | 2 | 1 | 2 |
| **Parietal lobe** | **Left** | **Supramarginal gyrus** | 0 | 0 | 1 | 2 | 1 | 2 |
| **Parietal lobe** | **Right** | **Supplementary motor cortex** | 0 | 0 | 1 | 2 | 2 | 3 |

**S9 Table 17.** Best regions for predicting True Positives (TP, i.e. true diagnosis of Autism) for Girls aged 10 to 15.

| | | | R42 | | D32 | | D70 | |
|---|---|---|---|---|---|---|---|---|
| | | | No comorb | With comorb | No comorb | With comorb | No comorb | With comorb |
| **temporal lobe** | **Right** | **Postcentral gyrus medial segment** | 0 | 0 | 2 | 2 | 2 | 2 |

**S9 Table 18.** Best regions for predicting True Positives (TP, i.e. true diagnosis of Autism) for Boys aged 15 to 20.

| | | | R42 | | D32 | | D70 | |
|---|---|---|---|---|---|---|---|---|
| | | | No comorb | With comorb | No comorb | With comorb | No comorb | With comorb |
| **Parietal lobe** | **Left** | **Parietal operculum** | 1 | 1 | 1 | 1 | 2 | 2 |

**S9 Table 19.** Best regions for predicting True Positives (TP, i.e. true diagnosis of Autism) for Girls aged 20 to 64.

**S10 - *Multi-site effect***

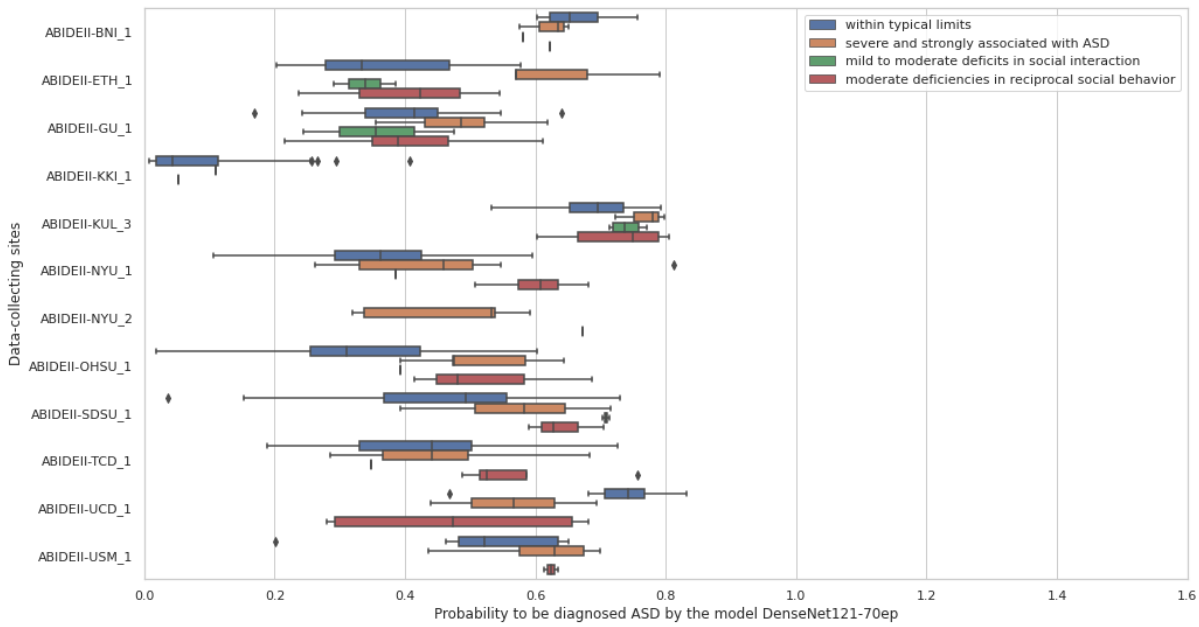


**S10 Fig 7.** Comparison of probabilities of Med3d-ResNet50-42ep and categories obtained from SRS T-scores for different sites

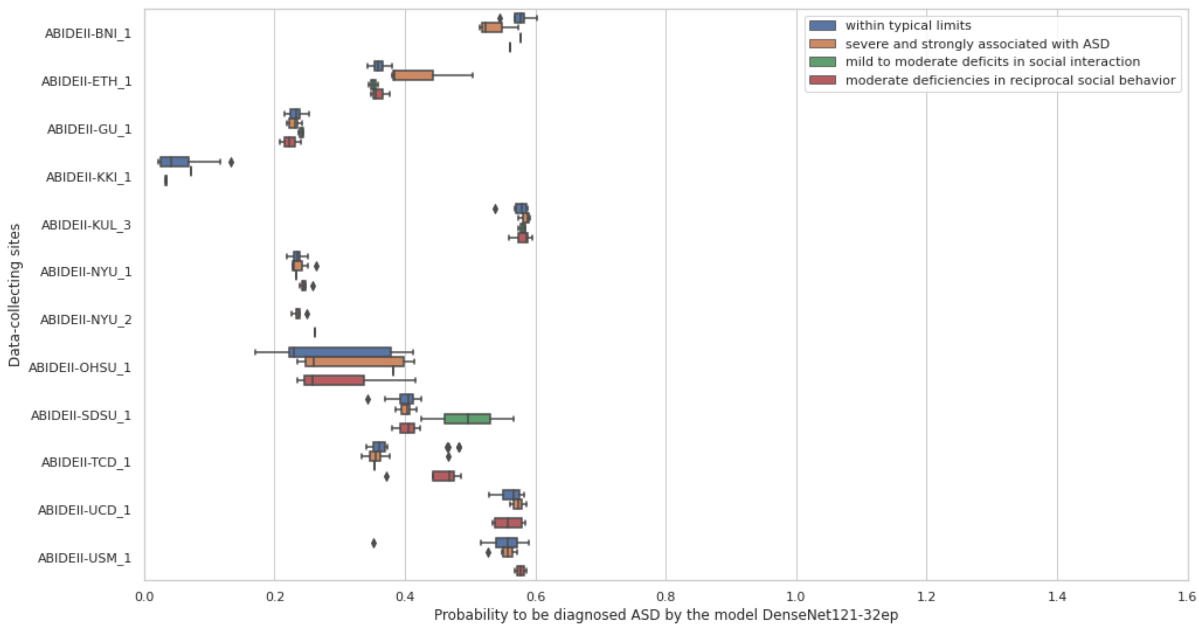


**S10 Fig 8.** Comparison of probabilities of DenseNet121-32ep and categories obtained from SRS T-scores for different sites

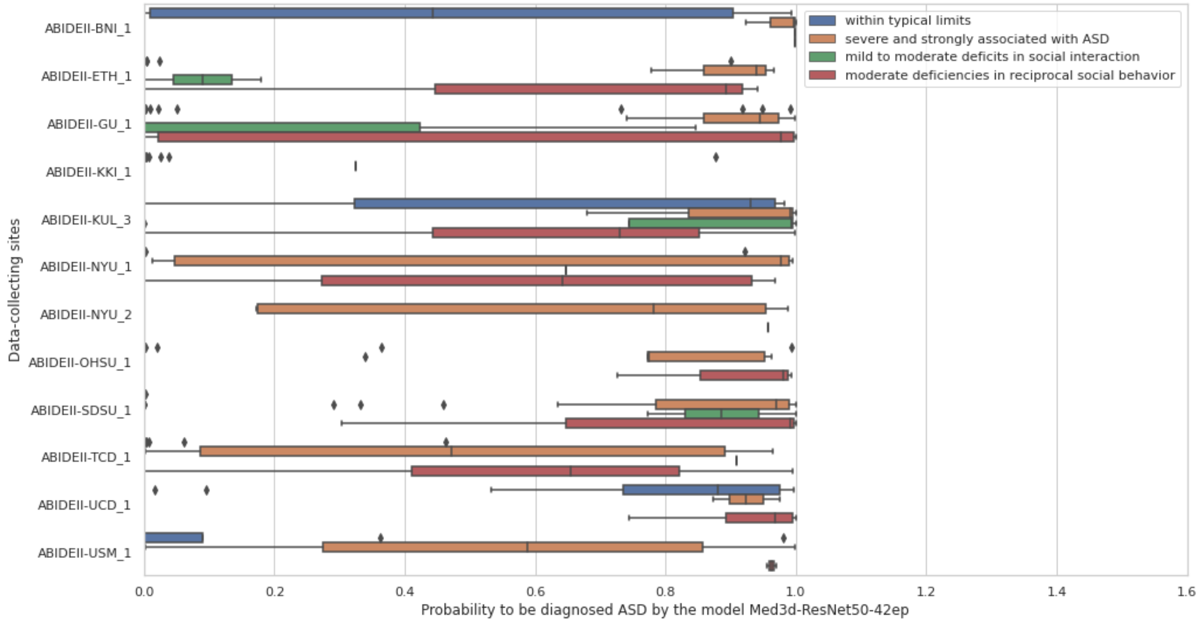


**S10 Fig 9.** Comparison of probabilities of DenseNet121-70ep and categories obtained from SRS T-scores for different sites

In **S10 Fig 7**, **S10 Fig 8**, and **S10 Fig 9**, we observe an inhomogeneous consistency of the distributions of probability scores between the different sites. Results in **S10 Table 20**, which displays the accuracy scores for every site in the whole dataset (training+validation+testing sets), confirm the multi-site effect already observed in **S10 Fig 7**, **S10 Fig 8**, and **S10 Fig 9**.

| | | | **Med3d-ResNet50-42ep** | | | **DenseNet-32ep** | | | **DenseNet-70ep** | | |
|---|---|---|---|---|---|---|---|---|---|---|---|
| | **site** | **n** | **Acc** | **Acc Autism** | **Acc TD** | **Acc** | **Acc Autism** | **Acc TD** | **Acc** | **Acc Autism** | **Acc TD** |
| **Training or validation set** | **ABIDEII BNI_1** | **9** | 100.0 | 100.0 | 100.0 | 77.8 | 100.0 | 0.0 | 77.8 | 100.0 | 0.0 |
| | **ABIDEII ETH_1** | **30** | 96.7 | 87.5 | 100.0 | 76.7 | 12.5 | 100.0 | 83.3 | 62.5 | 90.9 |
| | **ABIDEII GU_1** | **65** | 98.5 | 95.2 | 100.0 | 67.7 | 0.0 | 100.0 | 72.3 | 33.3 | 90.9 |

| | | | | | | | | | | |
|---|---|---|---|---|---|---|---|---|---|---|
| **ABIDEII IP_1** | **36** | 86.1 | 63.6 | 96.0 | 69.4 | 0.0 | 100.0 | 69.4 | 54.5 | 76.0 |
| **ABIDEII IU_1** | **33** | 93.9 | 93.8 | 94.1 | 54.5 | 50.0 | 58.8 | 57.6 | 87.5 | 29.4 |
| **ABIDEII KKI_1** | **123** | 98.4 | 0.0 | 99.2 | 99.2 | 0.0 | 100.0 | 99.2 | 0.0 | 100.0 |
| **ABIDEII KUL_3** | **20** | 75.0 | 75.0 | NaN | 100.0 | 100.0 | NaN | 100.0 | 100.0 | NaN |
| **ABIDEII NYU_1** | **41** | 85.4 | 57.1 | 100.0 | 65.9 | 0.0 | 100.0 | 75.6 | 42.9 | 92.6 |
| **ABIDEII NYU_2** | **6** | 66.7 | 66.7 | NaN | 0.0 | 0.0 | NaN | 66.7 | 66.7 | NaN |
| **ABIDEII OHSU_1** | **55** | 98.2 | 90.9 | 100.0 | 80.0 | 0.0 | 100.0 | 80.0 | 45.5 | 88.6 |
| **ABIDEII SDSU_1** | **55** | 89.1 | 80.6 | 100.0 | 45.5 | 3.2 | 100.0 | 69.1 | 80.6 | 54.2 |
| **ABIDEII TCD_1** | **36** | 80.6 | 56.2 | 100.0 | 55.6 | 0.0 | 100.0 | 52.8 | 31.2 | 70.0 |
| **ABIDEII USM_1** | **23** | 91.3 | 83.3 | 100.0 | 56.5 | 100.0 | 9.1 | 73.9 | 91.7 | 54.5 |
| **CALTECH** | **36** | 94.4 | 94.4 | 94.4 | 50.0 | 0.0 | 100.0 | 52.8 | 94.4 | 11.1 |
| **CMU** | **27** | 88.9 | 78.6 | 100.0 | 48.1 | 0.0 | 100.0 | 66.7 | 71.4 | 61.5 |
| **KKI** | **25** | 84.0 | 33.3 | 100.0 | 76.0 | 0.0 | 100.0 | 76.0 | 0.0 | 100.0 |
| **LEUVEN_2** | **31** | 83.9 | 61.5 | 100.0 | 41.9 | 100.0 | 0.0 | 67.7 | 84.6 | 55.6 |
| **MAX_MUN** | **33** | 87.9 | 69.2 | 100.0 | 60.6 | 0.0 | 100.0 | 51.5 | 23.1 | 70.0 |
| **NYU** | **124** | 91.9 | 68.8 | 100.0 | 74.2 | 0.0 | 100.0 | 69.4 | 31.2 | 82.6 |

| | | | | | | | | | | | |
|---|---|---|---|---|---|---|---|---|---|---|---|
| | **OHSU** | **22** | 77.3 | 63.6 | 90.9 | 50.0 | 0.0 | 100.0 | 59.1 | 90.9 | 27.3 |
| | **OLIN** | **19** | 63.2 | 41.7 | 100.0 | 63.2 | 100.0 | 0.0 | 63.2 | 100.0 | 0.0 |
| | **PITT** | **39** | 84.6 | 73.7 | 95.0 | 51.3 | 0.0 | 100.0 | 51.3 | 84.2 | 20.0 |
| | **SBL** | **29** | 75.9 | 50.0 | 100.0 | 58.6 | 85.7 | 33.3 | 48.3 | 92.9 | 6.7 |
| | **SDSU** | **12** | 100.0 | 100.0 | 100.0 | 83.3 | 0.0 | 100.0 | 75.0 | 100.0 | 70.0 |
| | **STANFORD** | **7** | 57.1 | 0.0 | 100.0 | 57.1 | 0.0 | 100.0 | 57.1 | 0.0 | 100.0 |
| | **TRINITY** | **42** | 85.7 | 70.0 | 100.0 | 52.4 | 0.0 | 100.0 | 50.0 | 40.0 | 59.1 |
| | **UCLA_1** | **52** | 86.5 | 81.2 | 95.0 | 61.5 | 100.0 | 0.0 | 65.4 | 87.5 | 30.0 |
| | **UCLA_2** | **15** | 93.3 | 100.0 | 88.9 | 46.7 | 100.0 | 11.1 | 53.3 | 100.0 | 22.2 |
| | **UM_1** | **72** | 90.3 | 74.1 | 100.0 | 62.5 | 0.0 | 100.0 | 66.7 | 22.2 | 93.3 |
| | **UM_2** | **31** | 93.5 | 83.3 | 100.0 | 61.3 | 0.0 | 100.0 | 77.4 | 66.7 | 84.2 |
| | **USM** | **66** | 87.9 | 83.7 | 95.7 | 63.6 | 95.3 | 4.3 | 62.1 | 93.0 | 4.3 |
| | **YALE** | **50** | 88.0 | 79.2 | 96.2 | 52.0 | 0.0 | 100.0 | 62.0 | 87.5 | 38.5 |
| **Testing set** | **LEUVEN_1** | **27** | 44.4 | 21.4 | 69.2 | 51.9 | 100.0 | 0.0 | 48.1 | 64.3 | 30.8 |
| | **ABIDEII EMC_1** | **18** | 72.2 | 50.0 | 78.6 | 77.8 | 0.0 | 100.0 | 38.9 | 100.0 | 21.4 |
| | **ABIDEII UCD_1** | **20** | 50.0 | 100.0 | 16.7 | 40.0 | 100.0 | 0.0 | 30.0 | 62.5 | 8.3 |

**S10 Table 20.** Comparing accuracy scores between data collection sites

For Med3d-ResNet50-42ep, the overall accuracy scores are between 44,4% - 100%, with 75% of the data-collecting sites having an accuracy higher than 78,9%, and an overall median accuracy of 87,9%. The sensitivity is between 0% - 100%, with a median of 74%. The specificity is between 16,7% - 100%, with a median of 100%.

For DenseNet121-32ep, the overall accuracy scores are between 0% - 100%, with 75% of the data-collecting sites having an accuracy higher than 51,6%, and an overall median accuracy of 60,6%. The sensitivity is between 0% - 100%, with a median of 0%. The specificity is between 0% - 100%, with a median of 100%.

For DenseNet121-70ep, the overall accuracy scores are between 30% - 100%, with 75% of the data-collecting sites having an accuracy higher than 53,1%, and an overall median accuracy of 66,7%. The sensitivity is between 0% - 100%, with a median of 71,4%. The specificity is between 0% - 100%, with a median of 55,6%.
